# Genome-Wide Association Study of Risk for Eosinophilic Granulomatosis with Polyangiitis

**DOI:** 10.1101/2025.11.07.25339770

**Authors:** Sung Chun, Parul H Kothari, Seung-Han Baek, Ahmad Samiei, Michael H Cho, Michael Wechsler, Peter A Merkel, Benjamin A Raby, Vasculitis Clinical Research Consortium

## Abstract

Eosinophilic granulomatosis with polyangiitis (EGPA) is an anti-neutrophil cytoplasmic antibody-associated vasculitis characterized by the manifestation of asthma and eosinophilia in the early phases. Currently, it is not well understood how underlying genetic risk factors affect EGPA and its comorbidity. To address this question, we aim to identify novel genetic associations with EGPA and investigating their pleiotropy with molecular traits. We conducted a genome-wide association study (GWAS) in a discovery cohort combining two independent studies by meta-analysis. The combined study generated the largest GWAS of EGPA to date (829 cases and 9,586 controls). We performed replication in an independent cohort from European ancestry (99 cases and 1,680 controls). In current study, we identified and formally replicated a novel genome-wide significant association with EGPA in *SSH2* locus. In the full meta-analysis combining our discovery and validation cohort, we also identified an additional genome-wide significant association in *RUNX1* locus. We found significant evidence that the EGPA association in *SSH2* locus colocalizes with *trans*-Quantitative Trait Loci (QTLs) affecting DNA methylation of CpG sites in *IL5RA* (a biological target of anti-IL5 therapy for EGPA) and in *LYN* (a tyrosine kinase interacting with *IL5RA*), which are also observed in asthmatics. Our findings provide new targets for future functional studies to elucidate pathogenesis of EGPA.

## INTRODUCTION

Eosinophilic granulomatosis with polyangiitis (EGPA), formally known as Churg Strauss syndrome, is an anti-neutrophil cytoplasmic antibody (ANCA)-associated vasculitis characterized by asthma and eosinophilia^1,2^. In the prodromal phase, the majority of EGPA patients (95-100%) manifest with asthma^3^. Over time, the disease progresses toward the eosinophilic phase, marked by the elevated eosinophils in blood and tissues. In the vasculitic phase, eosinophil activation leads to granulomatous inflammation and systemic necrotizing vasculitis of small-to-medium vessels.

EGPA remains to be a complex genetic disorder with unclear etiology. Genetic studies^4,5^ have been challenging due to the extremely low prevalence (2 – 38 cases per million)^6^. However, a recent genome-wide association study conducted by European Vasculitis Genetics Consortium (EVGC) began to shed light on the genetic architecture of EGPA^7^. Despite the rarity, EGPA is surprising polygenic with eleven common susceptibility loci identified, four of which were formally replicated in an independent cohort. Several EGPA association peaks showed the genetic heterogeneity by MPO+ ANCA status in the patients. Further, extensive pleiotropy was observed between EGPA and related traits such as asthma and eosinophil count.

Here, we expand the genome-wide association study of EGPA to a discovery cohort with a total of 829 cases and 9,586 controls from European ancestry by meta-analyzing Vasculitis Clinical Research Consortium (VCRC) with EVGC GWAS at the summary-statistics level. This was followed by replication in a validation cohort consisted of 99 cases and 1,680 controls from European ancestry. Finally, to improve our understanding of EGPA pathogenesis, we investigated transcriptomic and epigenetic variations that underlie EGPA association peaks.

## RESULTS

The workflow diagram in **Figure 1** summarizes our three-stage GWAS strategy. We started by carrying out a GWAS in 295 EGPA cases from the Vasculitis Clinical Research Consortium (VCRC) and 2,898 matching controls, all from European ancestry. These subjects form part of our discovery cohort. The control subjects were randomly sampled from healthy individuals from UK Biobank^8^, excluding individuals with history of EGPA, asthma and other respiratory disease, common autoimmune disorders, or vasculitis-related conditions. While UK Biobank subjects were primarily of British White ancestry, our EGPA patients were sampled more widely across European ancestry. Therefore, to avoid potential population stratification, we sampled a subset of control subjects matching the fine-scale ancestry of cases in the top two principal component (PC) space. Then, we tested for the genetic association under a logistic regression model, additionally adjusting for the genetic ancestry by incorporating the top ten PCs as covariates. The resulting association statistics showed a low genomic inflation (α_GC_ = 1.022), indicating that the population structure was well-controlled.

**Figure 1.**
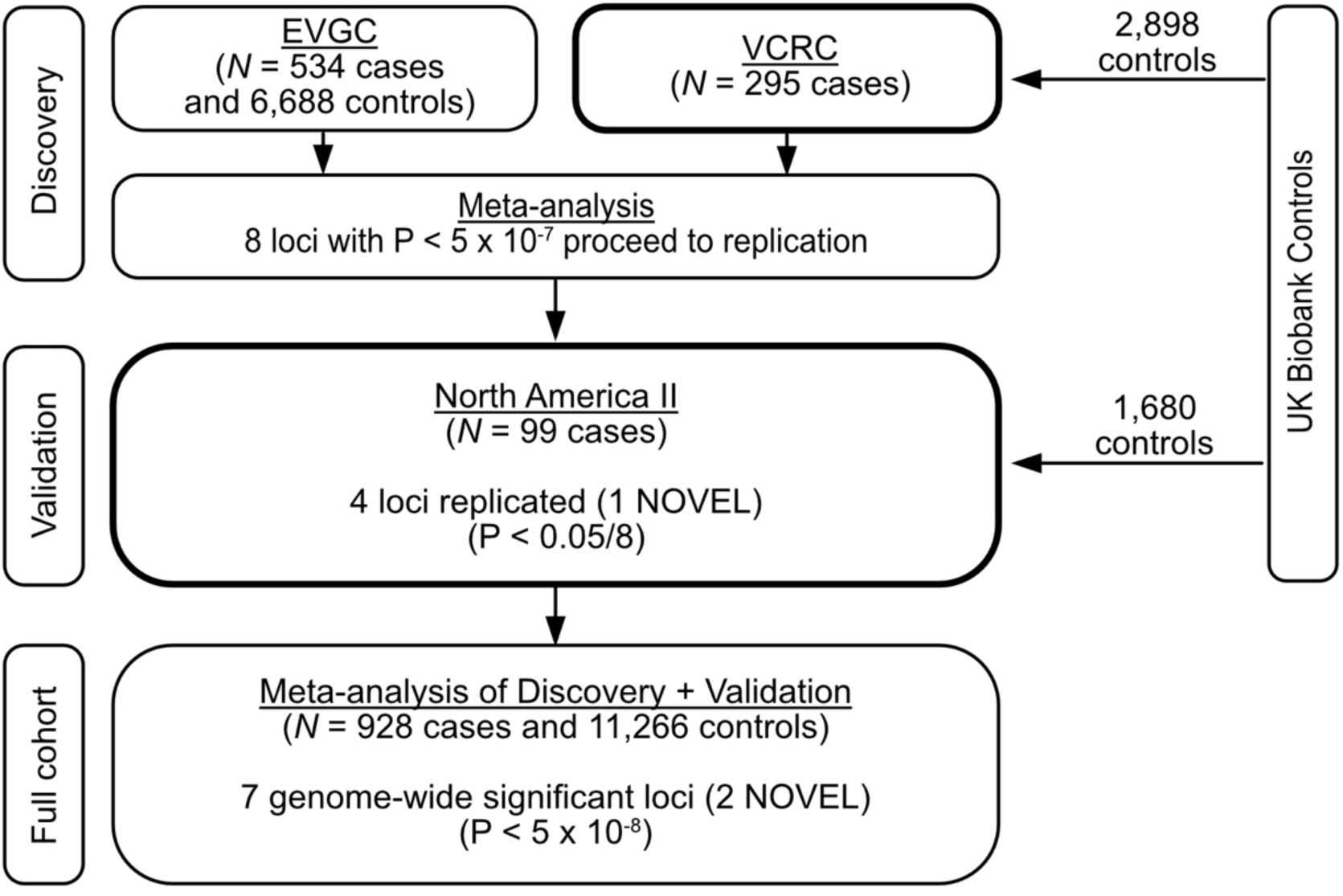
Overview of three-stage GWAS analysis approach. EGPA patients from the Vasculitis Clinical Research Consortium (VCRC) were combined with matching healthy controls from the UK Biobank. Genetic association results from VCRC/UK Biobank were then combined with European Vasculitis Genetics Consortium (EVGC) GWAS by meta-analysis at the summary-statistics level. A total of eight loci satisfying the discovery meta-analysis P-value < 5 x 10^−8^ proceeded to formal replication in an independent validation cohort (North America II). Finally, full meta-analysis combining discovery and validation cohorts was performed to achieve higher statistical power.

We confirmed significant genetic associations with EGPA for 4 out of the 11 previously reported loci by the European Vasculitis Genetics Consortium (EVGC)^7^ (**Supplementary Table 1**). All four loci that were genome-wide significant and formally validated in EVGC were also replicated in our cohort with consistent directions of effects (*BCL2L11*, *TSLP*, *HLA-DQ*, and 10q14). Also, we replicate the variation of genetic effect by ANCA status in *HLA-DQ* locus. As in the EVGC GWAS, the genetic association of *HLA-DQ* was significantly larger for MPO+ ANCA-positive subtype (OR = 2.55, 95% CI = [1.83 – 3.56]) compared to ANCA-negative subset (OR = 1.07, 95% CI = [0.84 – 1.37]). On the other hand, we failed to replicate three other loci which had been identified by genetic analyses stratified by ANCA status in the EVGC study (*GPA33*, *HLA* and 12q21). The lack of replication can be due to the limited sample size of VCRC in each ANCA stratum (81 MPO+ ANCA+ and 190 ANCA-EGPA patients), modest linkage disequilibrium (LD) of our proxy SNP to the lead SNP in case of *HLA* (r^2^ = 0.65), or non-replication in the original study. Lastly, the EVGC study identified four additional EGPA associations by leveraging the pleiotropy between EGPA and related traits, such as asthma and blood eosinophil count. We could not replicate these signals at the reported lead SNPs with the current VCRC sample size. Applying a pleiotropy-based genetic association test to a small underpowered cohort is known to be susceptible to overestimation of effect sizes due to the winner’s curse^9^. Indeed, we find that the magnitude of estimated effect sizes of these loci was substantially smaller in VCRC than was previously reported in EVGC.

To increase statistical power, we conducted a meta-analysis of GWAS data across VCRC and EVGC at a summary statistics level (**Figure 1**). After the harmonizing two GWAS datasets, we carried out a meta-analysis between two datasets for 7,254,833 shared SNPs using Stouffer’s method (a total of 829 EGPA cases and 9,586 controls). The meta-analysis p-values in our combined discovery cohorts showed a well-controlled genomic inflation (α_GC_ = 1.017). A total of 507 SNPs in five loci reached genome-wide significant p-values (P < 5 x 10^−8^). The five loci include four previously known EGPA loci and *IRF1/IL5*. *IRF1/IL5* locus was originally identified based on its pleiotropy with EGPA-related traits previously^7^ and is now genome-wide significant in the meta-analysis combining EVGC and VCRC (P = 4.2 x 10^−8^). Further, we found 727 SNPs in eight loci with suggestive p-values (P < 5 x 10^−7^). Considering one lead SNP for each locus, eight lead SNPs surpassing the suggestive p-value threshold were taken forward for formal replication in a separate validation cohort of 99 North American EGPA cases of European ancestry (North America II). Matching controls (1,680 non-overlapping healthy subjects) were sampled from the UK Biobank, similarly as for VCRC cohort. Of the eight SNPs from the discovery stage, only one was formally replicated after the Bonferroni correction (P = 1.9 x 10^−3^), showing concordant directions of effect across all three cohorts (**Supplementary Table 2**). This association signal is novel and located in the *SSH2* gene (which encode Slingshot Protein Phosphatase 2; SSH2). Given the extreme rarity of EGPA in population, our validation cohort is limited in the sample size and statistical power. As such, other EGPA loci, even if they are previously known, did not pass the formal multiple testing threshold in our validation cohort.

To further increase statistical power and fine-map association signals, we conducted a three-way meta-analysis combining VCRC, EVGC and North America II (a total of 928 cases and 11,266 controls). The meta-analysis showed with a well-controlled genomic inflation (α_GC_ = 1.056). Overall, seven loci passed the genome-wide significance threshold (P < 5 x 10^−8^), of which five are previously known loci (**Figure 2**, **Supplementary Table 3**). The GWAS peak of *SSH2* locus was localized to a 500 kb region around the lead SNP in the first intron of *SSH2* (**Figure 3A**). In addition, we identified another novel genome-wide significant association in the upstream non-coding region of *RUNX1* gene (P = 9.5 x 10^−9^), which encodes RUNX family transcription factor 1 (**Figure 3B**). For both *SSH2* and *RUNX1*, the directions of effects were concordant across all three cohorts (**Table 1**). Unlike *HLA-DQ*, we did not observe a significant difference in effect size by ANCA status in both novel loci. In VCRC, the OR of *SSH2* locus was 1.63 (95% CI = [1.10 – 2.41]) in MPO+ ANCA+ compared to 1.51 (95% CI = [1.16 – 1.98]) in ANCA-negative group. The OR of *RUNX1* locus was 0.82 (95% CI = [0.58 – 1.16]) in MPO+ ANCA+ compared to 0.80 (95% CI = [0.63 – 1.01]) in ANCA-negative subgroup.

**Figure 2.**
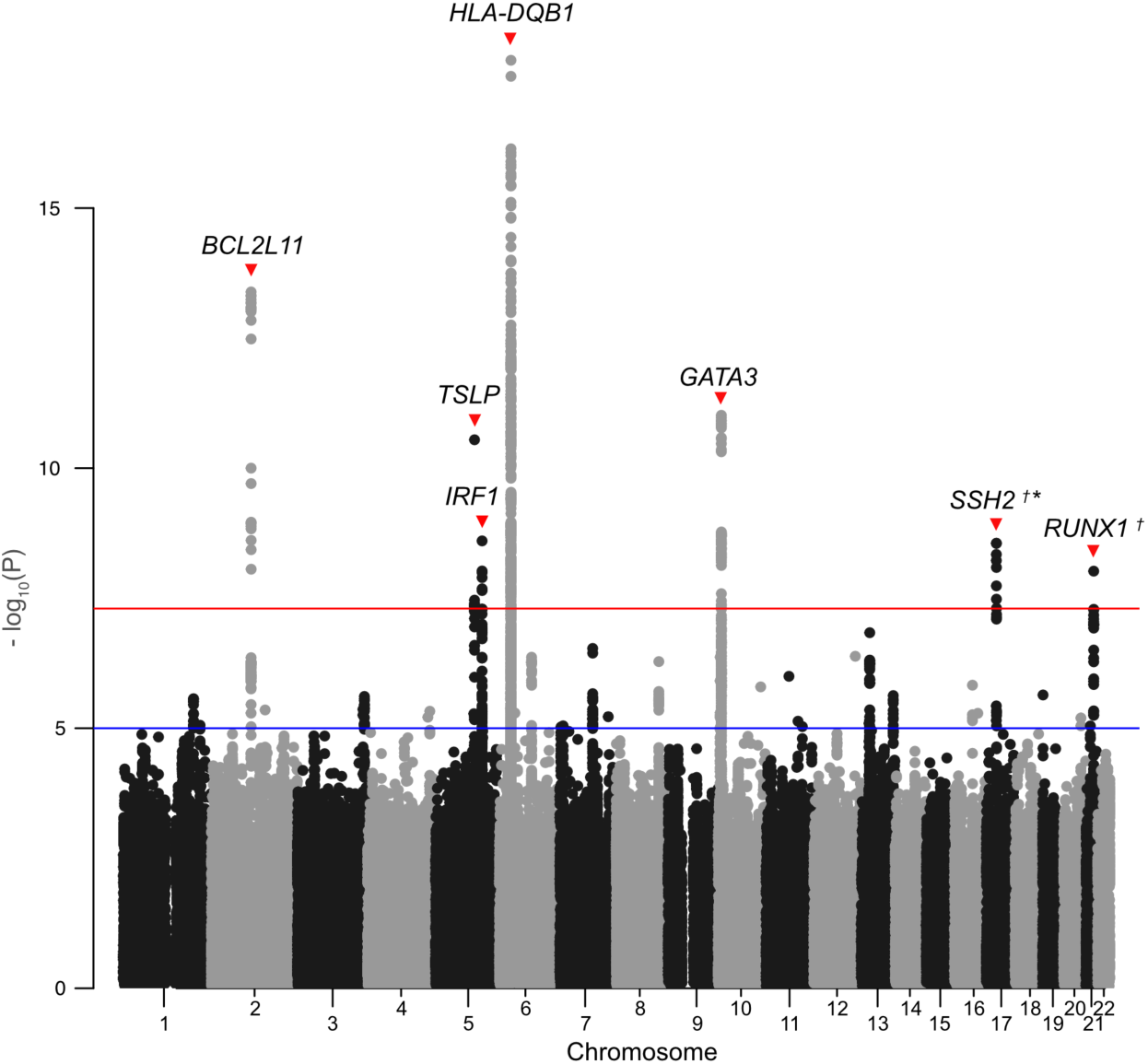
Manhattan plot of three-way EGPA GWAS meta-analysis across VCRC, EGVC and North America II. One SNP in chromosome 11 have inconsistent directions of effect across the three cohorts. We replicate five previously known EGPA-associated loci. The nearest genes for each lead SNP are indicated for each locus. *The association in *SSH2* locus was formally replicated in an independent validation cohort. †We identified two novel genome-wide significant associations to EGPA in *SSH2* and *RUNX1* loci.

**Figure 3.**
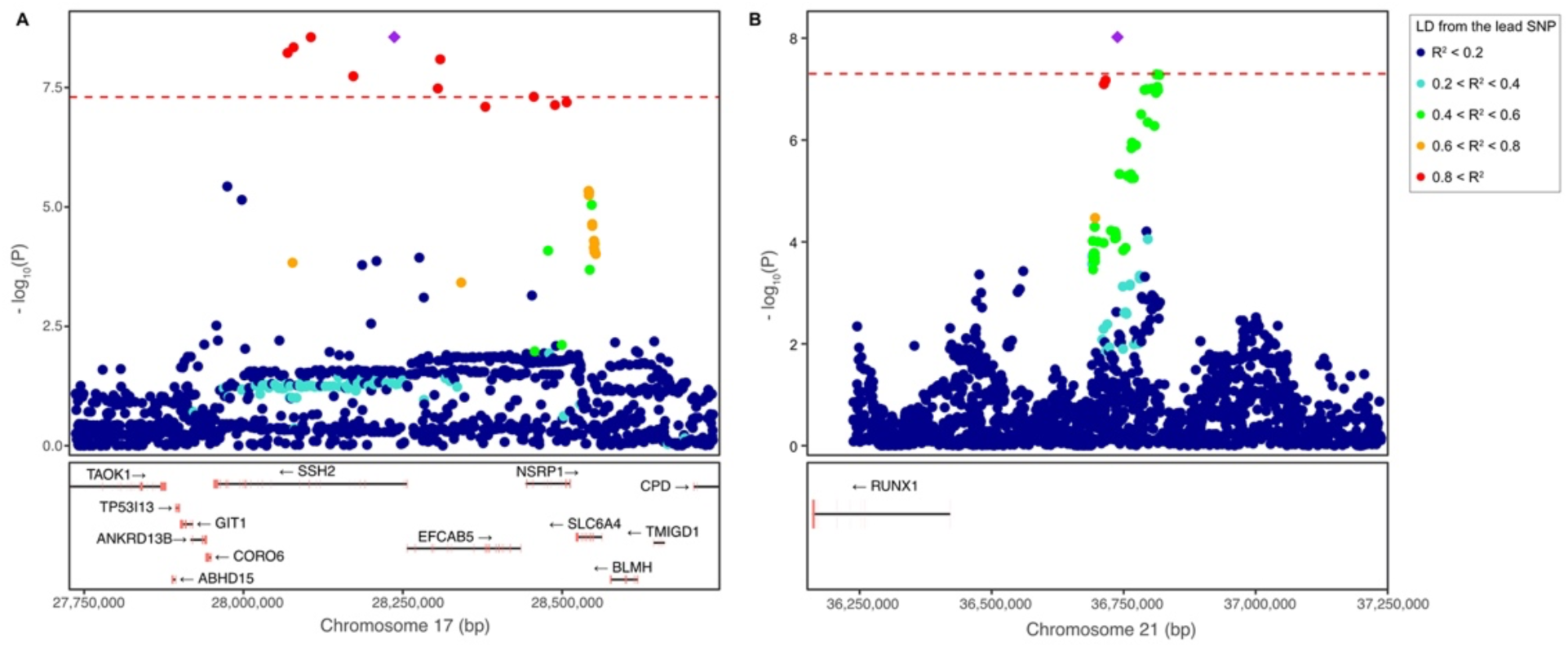
Region plots of two novel genome-wide significant eosinophilic granulomatosis with polyangiitis (EGPA)-associated loci identified from the full meta-analysis combining all three cohorts. Each point represents a SNP with the chromosomal coordinate on the x axis and the p-value of association on the y axis. Purple diamond indicates the lead SNP with the most significant p-value. Points were colored based on the linkage disequilibrium (r^2^) of each SNP from the lead SNP. Dashed red line indicates the genome-wide significance level (5 x 10^−8^). **(A)** EGPA association peak in *SSH2* locus. **(B)** EGPA association peak in *RUNX1* locus.

A prior study reported the extensive pleiotropy between EGPA and other traits, notably asthma and eosinophil count^7^. Further, we noted that the EGPA lead SNP in *RUNX1* locus (rs8133843) is in a strong LD with the lead SNP for a genome-wide significant association for rheumatoid arthritis (rs9979383, r^2^ = 0.90 in the 1000 Genomes Project Europeans)^10^. To more carefully examine if the observed locus-level pleiotropy is indeed driven by the same causal genetic effect, we compared the fine-resolution causal likelihoods between a pair of GWAS data while accounting for the underlying LD structure using a colocalization test. Specifically, for each genome-wide significant EGPA-associated locus, we applied Joint Likelihood Mapping (JLIM)^11^ to genetic association data from EGPA and other traits, namely, adult-onset asthma^12^, rheumatoid arthritis^13^ and the variation of peripheral eosinophil counts in healthy population^14^ (**Supplementary Figure 3**, **Supplementary Table 4**). We found that the EGPA association peak in *IRF1/IL5* locus is driven by a distinct causal variant from asthma and eosinophil count in a fine resolution (JLIM P = 1.0 for both) although this locus was originally identified by a locus-level pleiotropy test (conditional false discovery rate test). In contrast, the EGPA association in *RUNX1* showed a significant colocalization not only with rheumatoid arthritis (JLIM P < 1.0 x 10^−6^) but also with asthma (JLIM P = 1 x 10^−5^) and peripheral eosinophil count (JLIM P < 1 x 10^−6^). The risk allele for EGPA (A) was associated with high risk for asthma and rheumatoid arthritis and increased eosinophil counts in blood.

Lead SNPs of all currently known and novel genome-wide significant EGPA association peaks land in non-coding regulatory regions. To predict likely causal genes underlying these EGPA associations, we examined using JLIM whether the variation in transcriptional levels of any protein-coding gene is driven by the same genetic variants as EGPA GWAS peaks. Specifically, we tested for the colocalization between EGPA GWAS and expression quantitative trait locus (eQTL) from whole bloods and immune cell populations such as neutrophils, monocytes and naïve CD4+ T cells from both asthmatic and healthy individuals^15–17^. The eQTL analysis revealed significant evidence of colocalization of EGPA association and *SSH2* eQTL in whole bloods from the Estonian Biobank (JLIM P = 1.9 x 10^−4^; FDR = 0.012) (**Supplementary Table 5**). The EGPA risk allele of lead SNP (rs8073292) is associated with a lower expression of *SSH2* in whole bloods in Estonians (**Supplementary Figure 2**). Knockdown of *SSH2* is known to cause the reduction of polarization and chemotaxis of neutrophils in mice^18^. In this locus, we also observed significant colocalization of EGPA association with the cytokine receptor like factor 3 (*CRLF3*) eQTL in naïve CD4+ T cells from asthmatic children (JLIM P = 7.3 x 10^−4^, FDR = 0.025). *CRLF3* is an orphan cytokine receptor involved in a negative regulation of anti-viral immunity in zebrafish^19^. The risk allele of rs8073292 is associated with a lower expression of *CRLF3*, thus with an increased anti-viral immune reaction.

EGPA-associated genetic variants may affect disease risks epigenetically by dysregulating DNA methylation^20^. Gene-specific differential DNA methylation has been implicated in ANCA-associated vasculitis^21,22^, other autoimmune disorders^23–26^ and asthma^27^. Loss or gain of DNA methylation at CpG dinucleotides can alter gene expression by dysregulating the accessibility of non-coding cis-regulatory elements. Using JLIM, we investigated whether any of genome-wide significant EGPA association peaks colocalize with whole blood methylation QTL (mQTL) from 110 Mexican asthmatic children in Asthma BRIDGE ^28,29^. Although none of these asthmatic children had EGPA, we hypothesized that EGPA and asthma might share common epigenetic regulatory pathways, and if so, we might be able to observe them by the colocalization of EGPA-associated variants with blood mQTL in asthmatics. Indeed, EGPA association in *SSH2* significantly colocalize with the *cis*-mQTL of an intronic CpG site in nearby gene, *EFCAB5*, which encodes EF-Hand Calcium Binding Domain 5 (JLIM P = 1.0 x 10^−4^, FDR = 0.004, **Figure 4A**, **Supplementary Table 6**). Although its function is not well-understood, *EFCAB5* is most highly expressed in eosinophils and basophils compared to other blood cell types^30^. The EGPA risk allele (A) is associated with hypomethylation of the CpG site (cg24184684) in *cis*, thus likely to lead to derepression of expression in EGPA.

**Figure 4.**
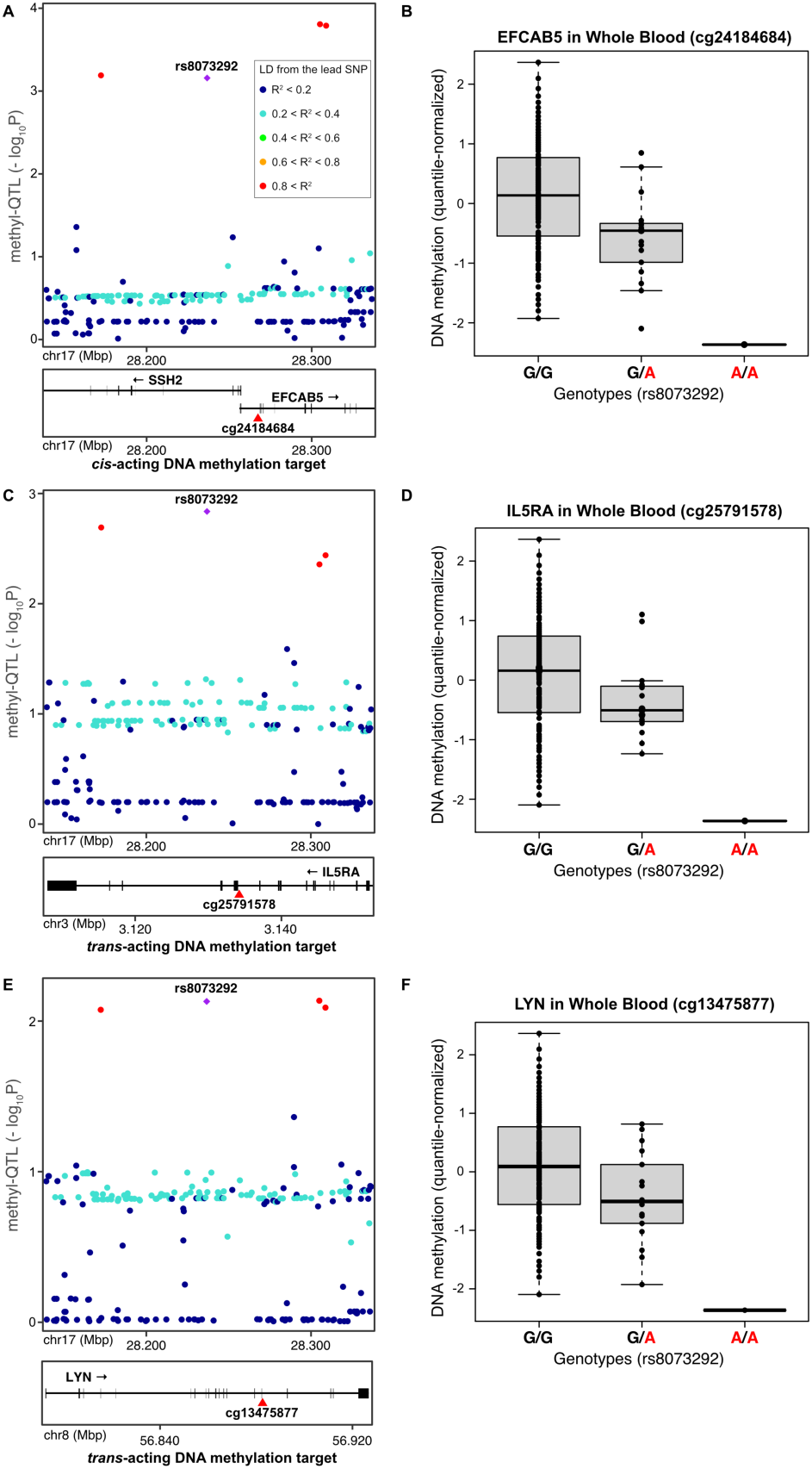
*cis*– and *trans*-acting DNA methylation quantitative trait loci (mQTL) colocalizing with the eosinophilic granulomatosis with polyangiitis (EGPA) association peak in *SSH2* locus. EGAP association peak in chromosome 17 colocalizes with. (A) *EFCAB5 cis*-mQTL, (C) *IL5RA trans*-mQTL, and (E) *LYN trans*-mQTL in whole blood DNA samples. EGPA risk allele is associated with hypomethylation of CpG site in (B) *EFCAB5* in chromosome 17, (D) *IL5RA* in chromosome 3, and (F) *LYN* in chromosome 8. In the region plots (left), each point represents a SNP with the chromosomal coordinate on the x axis and the p-value of mQTL association on the y axis. Purple diamond indicates the lead SNP for EGPA association (rs8073292). Points were colored based on the linkage disequilibrium (LD) of each SNP from the lead SNP. EGPA lead SNP and close LD neighbors (r^2^ > 0.8, marked in red) drive mQTL signals, consistent with the shared genetic effect between EGPA and mQTL. Red triangle represents the location of CpG dinucleotide. In the box plots (right), each point represents a subject with the genotype at rs8073292 on the x axis and DNA methylation levels on the y axis. DNA methylation levels were quantile-normalized. A is the risk allele for EGPA at rs8073292. DNA methylation data are from asthmatic children in Mexico obtained through Asthma BRIDGE consortium (N = 110).

EGPA-associated variants might lead to dysregulated DNA methylation of CpG sites in *trans* as well. Since our sample size of methylation dataset is limited for comprehensive genome-wide *trans*-mQTL scan, we limited our search to seven genes that affect blood cytokine levels, are differentially methylated, and show strongest evidence of differential mRNA expression in asthma (*CHUK, IL1R2, IL1RN, IL5RA, IRF7, LYN,* and *SOCS2*)^31^. Interestingly, EGPA association in the *SSH2* locus showed significant evidence of colocalization with a *trans-* mQTL on *IL5RA*, which encodes interleukin-5 receptor subunit alpha in chromosome 3 (JLIM P = 9.0 x 10^−4^, FDR = 0.010, **Figure 4B**, **Supplementary Table 7**). IL5RA is the biological target of Benralizumab (IL5RA antagonist), one of the most effective treatments currently available for EGPA^32^. Additionally, EGPA association in *SSH2* significantly colocalize with a *trans-*mQTL on *LYN*, which encodes Lyn tyrosine kinase in chromosome 8 (JLIM P = 2.5 x 10^−3^, FDR = 0.014, **Supplementary Table 7**). LYN kinase catalyzes the IL5-stiumulated tyrosine phosphorylation of IL5RA in eosinophils and plays a key role in eosinophil differentiation^33^. For both *IL5RA* and *LYN*, the EGPA risk allele (A) is associated with hypomethylation of CpG sites, consistent with the possibility of aberrant loss of silencing of IL5RA pathway in eosinophils.

## DISCUSSION

We undertook the largest GWAS of EGPA susceptibility to date and identified two novel signals of association in *SSH2* and *RUNX1* loci. The GWAS signal in the *SSH2* locus was formally replicated in an independent validation cohort. The *RUNX1* locus was found genome-wide significant in the meta-analysis combining our discovery and validation cohorts. None of our novel loci showed a significant heterogeneity of genetic effect by ANCA status in VCRC cohort.

The lead SNP of EGPA association in *SSH2* locus is located in the first intron of *SSH2*. Supporting the possibility that the EGPA association in this locus affects the disease risk by the regulatory variation of *SSH2* expression, we indeed detect the significant colocalization of the EGPA GWAS peak with *SSH2* eQTL in whole blood from Estonians. *SSH2* is known to affect polarization and chemotaxis of neutrophils^18^. However, evidence supporting the causal role of *SSH2* is conflicting. We did not observe significant colocalization of *SSH2* eQTL in neutrophils from heathy individuals (BLUEPRINT)^15^ or in whole bloods from asthmatic children (Asthma BRIDGE)^17^. Rather, in Asthma BRIDGE, we observed significant colocalization of eQTL of another gene, *CRLF3*, in naïve CD4+ T cells. The ligand of *CRLF3* cytokine receptor is unknown. However, in zebrafish, the ortholog plays a role in the negative regulation of anti-viral immunity^19^. Prior studies suggest that this gene plays a key role in hematopoiesis across zebrafish^34^ to mice (red blood cell and platelet maturation)^35^ and human (lymphoid cell counts)^36^.

Our results suggest that the EGPA risk allele in *SSH2* locus may involve DNA hypomethylation of CpG sites in *cis* and *trans*. The EGPA association peak significantly colocalizes with *cis-*mQTL affecting a CpG site in nearby *EFCAB5* and *trans-*mQTL regulating CpG sites in *IL5RA* and *LYN* in two different chromosomes. IL5RA is the biological target of current treatment for EGPA (Benralizumab)^32^, and LYN is a Src-family tyrosine kinase that binds and phosphorylates IL5RA under IL-5 in eosinophils^33^. Src-family kinase inhibitor (Dasatinib) has been developed to treat chronic myeloid leukemia but not been tested for the use in EGPA.

While the direction of EGPA risk allele is consistent with the increased eosinophilic inflammation observed in EGPA, colocalization tests cannot distinguish the causality from the horizontal pleiotropy. Thus, further mechanistic studies are necessary to formally establish the causal chain linking the EGPA-associated genetic variant to epigenetic changes. Another limitation of current study is that our DNA methylation data are from whole bloods. As such, we cannot completely rule out the possibility that the observed whole blood mQTL might simply reflect the variation of eosinophil fractions in peripheral bloods rather than different methylation states of eosinophils. Nonetheless, EGPA-associated variant in *SSH2* is not associated with the variation of eosinophil counts in blood (eosinophil count GWAS P = 0.81 at rs8073292)^14^, thus is unlikely to drive mQTL effect entirely through cell fraction changes. In contrast, EGPA GWAS peaks in *BCL2L11* and *RUNX1* loci show strong evidence of pleiotropy to eosinophil count GWAS (JLIM P = 2.2 x 10^−3^ and 1.0 x 10^−6^, respectively; **Supplementary Table 4**), yet they do not colocalize with any cis-or trans-mQTL (**Supplementary Tables 6-7**).

Notably, *SSH2* locus implicates the sharing of IL-5 genetic pathway between EGPA and asthma, which might explain why EGPA starts with asthma manifestation in the prodromal phase. Yet the lead EGPA-associated SNP in this locus (rs8073292) does not show pleiotropy to asthma (asthma GWAS P = 0.12)^12^. Here, the comorbidity seems to be induced by epigenetic modulation of downstream genes. On the other hand, *RUNX1* locus shows robust evidence of pleiotropy across asthma, eosinophil count, and rheumatoid arthritis (**Supplementary Table 4**). Interestingly, *RUNX1* (along with *RUNX3)* has shown to regulate the proliferation of group 2 innate lymphoid cells (ILC2s) and ILC2-mediate TH2 cytokine production during allergic airway inflammation^37^. Further, circulating ILC2s in blood have been implicated to rheumatoid arthritis^38^.

In summary, we report new biological insights into EGPA susceptibility and demonstrate that further genetic and epigenetic studies to understand the common mechanistic pathways underlying EGPA and asthma are needed.

## METHODS

### Study Cohorts

For the discovery cohort, a total of 380 EGPA patients were recruited across North America through the Vasculitis Clinical Research Consortium (VCRC). All VCRC DNA samples pass the quality control (QC) standard. For the validation cohort, a total of 129 EGPA patients were recruited across North America (North America II), independently from VCRC. After excluding 22 individuals with poor sample quality, 107 EGPA patients were available for the validation.

The study was conducted under an appropriate institutional review board (IRB) and ethics approval (IRB Protocol #: Partners Healthcare 2001P001765).

### Genotyping and quality control

DNA samples of all EGPA patients from both VCRC and North America II were genotyped with Affymetrix Axiom Biobank 1 microarrays according to the manufacture’s protocol. Genotyping of all samples was conducted by the Center for Applied Genomics at the Children’s Hospital of Philadelphia.

Standard QC practices were applied to genotyping data for VCRC samples. Specifically, four samples were excluded for the sex mismatch. Seven subjects with the missing rate > 5% were filtered out for poor genotyping quality. Eighteen subjects with a too high or too low heterozygosity (i.e., > 0.05 or < –0.05) and 23 subjects with non-European ancestry identified by the principal component analysis (PCA) were also excluded.

The identical QC criteria were applied to North America II. Four samples with the extreme heterozygosity were filtered out at the same heterozygosity cutoff. Eight individuals with non-European ancestry were also filtered out.

The heterozygosity calculation and genetic ancestry analysis were conducted using Plink. First, rare variants with minor allele frequencies (MAF) < 0.01 were removed. Remaining variants were further pruned to reduce the linkage disequilibrium (LD) between variants in windows of 100kb at the step of two variants and r^2^ threshold of 0.1. The threshold for extreme heterozygosity was selected at the point where the distribution of heterozygosity becomes discontinuous. Second, the genetic ancestry analysis was done on the genetic dataset of common variants (MAF > 0.05) that were similarly LD-pruned. Samples with non-European ancestry were identified by overlaying the genetic data of EGPA patients into the PCA space constructed with the 1000 Genome Project reference panel samples (phase 3 version 5).

Genetic relationship between EGPA patients were identified with KING in an identity-by-descent analysis. Fifteen duplicated samples were found across VCRC and North America II and were removed from VCRC. No related individuals (up to third degree relationship) were identified within or across these cohorts.

### Matching controls

A total of 214,400 control subjects were sampled from the UK Biobank^8^ by applying the following criteria. First, subjects with any EGPA-related condition were removed exhaustively from the control. Although EGPA is extremely rare, extensive pleiotropy has been observed between EGPA and other related conditions for many EGPA-associated variants^7^. For this, we excluded all subjects with a history of eosinophilia, any of the 24 most common autoimmune disorders, or any respiratory disorders. The medical history was scanned in the fields of self-reported illness, in-patient diagnosis, and death registry records. The details of control selection algorithm are described in **Supplementary Table 8**. Second, withdrawn subjects and those indicated for any QC issues were also excluded. The QC exclusion criteria include the sex mismatch, sex chromosome aneuploidy, outliers of the heterozygosity or genotyping missing rate, and exclusion from the genetic kinship analysis. Third, self-reported non-white participants were filtered out from the control. Lastly, to ensure that all control subjects are unrelated to each other, for each genetically inferred pairwise kinship, only one was kept randomly, eliminating the other.

Next, the selected healthy control subjects were further subsampled to match the fine-scale genetic ancestry of our case samples. In the UK Biobank, White British subjects constitute 87% of all self-reported Whites whereas our EGPA patients represent more diverse groups of European ancestry. To account for the fine-scale ancestry, the following down-sampling procedure was conducted on UK Biobank control samples, respectively for VCRC and North America II case populations. First, out of the control subjects, White British individuals were randomly thinned to 1:4 to make the subsequent sampling procedure more efficient. Second, the PCA space is constructed using White EGPA patients only, and then, all UK Biobank control subjects were projected into the PCA space. Third, the two-dimension space of top two ancestry-PC axes were divided into 40 x 40 grids. Then, the density of EGPA cases *f*(*i, j*) was estimated for each bin *i* ∈ {1, …, 40} and *j* ∈ {1, …, 40 }, using the bivariate averaged shifted histogram (ASH) algorithm^39^. This step was necessary since the distribution of EGPA cases were too sparse in most of the bins given the current case sample counts. The bivariate ASH conducts polynomial kernel density estimation across the two-dimensional grids. On the other hand, the density distribution of controls *g*(*i*, *j*) was derived directly from the counts in each bin since the sample size is large enough. Next, we conduct the acceptance-rejection sampling to match the proposal distribution *g*(*i*, *j*) to the target distribution *f*(*i, j*). Specifically, for each control sample (which belong to the bin *i* and *j*), a random variable *U* was generated from the uniform distribution Unif(0, 1), and the sample was accepted into the control only if:

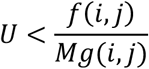

Here. *M* is a constant satisfying *f*(*i, j*) ≤ *Mg*(*i, j*) across all # and %. 6 is chosen to be as small as possible to increase the number of accepted controls. Here, the grid bins containing less than four samples (cases and controls combined) were masked out since they drive up 6. Note that for North America II, the 20 x 20 grid (instead of 40 x 40) was applied due to the smaller sample size.

In total, excluding seven cases present in the ancestry grids with sparsely distributed controls in the UK Biobank, 296 EGPA patients remained in VCRC. For controls, 2,898 subjects matching the fine-scale ancestry of cases were selected from the UK Biobank (**Supplementary Figure 4**). In North America II, no case was excluded in the ancestry grids, and a total of 99 EGPA patients and 1,680 matching controls were available (**Supplementary Figure 5**). No control subject was selected both for VCRC and North America II.

### Genotype imputation

Before imputation, the genotype data of cases and controls were QC-ed separately. For cases, variants with the genotyping rate < 95% or Hardy-Weinberg equilibrium P < 0.0001 were filtered out along with all InDels. For controls, variants with the genotyping rate < 95% and all InDels were excluded. Then, cases and controls were merged, keeping only the common set of genotyped markers. UK Biobank samples were genotyped with Affymetrix UK Biobank Axiom arrays, which have the similar set of markers as Axiom Biobank 1 arrays used for EGPA cases. Variants with the imbalance of genotype missingness between cases and controls were removed (P < 0.001) along with all rare variants with MAF < 1%. In total, 243,284 and 259,383 common SNPs passed the pre-imputation QC for VCRC/UK Biobank and North America II/UK Biobank cohorts, respectively.

The imputation was conducted against the Haplotype Reference Consortium (HRC release 1.1)^40^ using the Michigan Imputation Server^41^. Note that cases and controls were imputed together to minimize potential stratification. Excluding SNPs with MAF < 1% and INFO score < 0.4, a total of 7,551,085 imputed SNPs were available.

### Genetic association analysis

For each of 7,551,085 imputed SNPs, the genetic association was tested under the following logistic regression model:

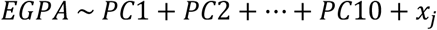

where *PC1, PC2, … PC10* are the top ten genetic ancestry PCs, and *x_j_* is the imputed genetic dosage of SNP j. The covariates were selected similarly as a prior GWAS in EGPA^7^. The genomic inflation was well controlled with λ*_GC_* = 1.022 for VCRC and λ*_GC_* = 1.012 for North America II, respectively.

For the meta-analysis, GWAS summary statistics were combined using Stouffer’s Z score method. Excluding 2,446 SNPs suspected of potential allele flip (the effect allele frequency difference > 0.2), a total of 7,254,833 SNPs were common between our study and EVGC. Since Z scores were not directly available for EVGC GWAS, the p-value *p_j_* of SNP *j* was converted to absolute Z score by the inverse cumulative distribution function Φ^−1^(⋅) and then was multiplied by the sign of estimated effect *β_j_* to obtain the signed Z score. The Z score was further adjusted by λ*_GC_* sine λ*_GC_* is relatively high for EVGC (1.047). Specifically, the Z score for EVGC was obtained as follows:

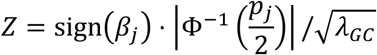

For VCRC and North America II, the original Z scores were used directly. The meta-analyzed Z scores were calculated using the following equation:

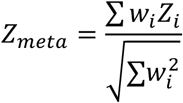

where Z*_i_* is the Z score and *w_i_* is the weight of GWAS cohort *i*. *w_i_* is set to 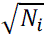, where *N_i_* is the effective sample size of cohort *i*. The final genomic inflation was well controlled for the discovery meta-analysis (λ*_GC_* = 1.017) and full three-way meta-analysis combining all cohorts (λ*_GC_* = 1.056).

### Gene expression quantitative trait loci (eQTL)

To identify candidate causal genes underlying EGPA GWAS peaks, *cis*-eQTL datasets of whole blood and immune cells were examined in various populations for which the reference linkage disequilibrium (LD) panels or in-sample LD were available. Specifically, for Estonian Biobank^16^, the genome-wide summary statistics for whole blood eQTL were downloaded from the eQTL catalogue^42^. The study subjects were from adult populations with European ancestry in Estonia (N = 471). The genome-wide summary statistics for BLUEPRINT^15^ were also obtained from the eQTL catalogue. The eQTL data for monocytes, neutrophils, and naïve T cells collected from a healthy adult population of European ancestry (N = 191, 196, and 196, respectively). Through the Asthma BRIDGE consortium, eQTL data were obtained for whole blood and T cells of asthmatic children. For other cell types in Asthma BRIDGE, the sample sizes were not sufficient for the eQTL analysis. Specifically, for whole blood, post-QC genetic (Illumina HumanHap550 v3) and transcriptomic (Illumina HumanHT-12 v4) data were obtained for 254 asthmatic children of European ancestry from the Childhood Asthma Management Program (CAMP) study^43^. For naïve CD4+ T cells, post-QC genetic (Affymetrix Genome-Wide Human SNP 6.0) and transcriptomic (Illumina HumanHT-12 v4) data were obtained for 68 asthmatic children of European ancestry from the Childhood Asthma Research and Education (CARE) network^44^. For each gene, the association between SNPs and expression levels of protein-coding genes was tested for all SNPs within +/− 1 Mbps from the transcription start site. Matrix-eQTL was used for the *cis*-eQTL association testing under the following linear regression model:

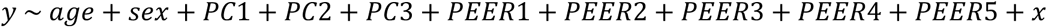

where *x* is the allelic dosage of tested SNP, *y* is the inverse normal transformed gene expression levels, *age* and *sex* are the age and sex of subject, *PC*1, *PC*2, and: *PC*3 are the top three genetic ancestry PCs, and: *PEER*1: *PEER*2, …,: *PEER*5 are the top five PEER factors accounting for batch effects in expression profiles^45^.

### DNA methylation quantitative trait loci (mQTL)

To test whether an EGPA-associated variant affects DNA methylation levels, *cis-* and *trans-*mQTL data were generated for whole blood samples through the Asthma BRIDGE consortium. Specifically, post-QC genetic (Illumina HumanHap550 v3) and DNA methylation (Illumina HumanMethylation 450K) data were obtained for 110 asthmatic children from the Mexico City Childhood Asthma Study (MCCAS) study^28,29^. For other cell types, the sample sizes available through Asthma BRIDGE were too limited for the mQTL analysis. For each CpG methylation site, the association between SNPs and DNA methylation levels was tested using Plink under the following linear regression model:

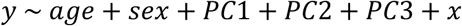

where *x* is the allelic dosage of tested SNP, *age* and *sex* are the age and sex of subject, and *PC*1, *PC*2, and *PC*3 are the top three genetic ancestry PCs. *y* is the inverse normal transformed DNA methylation betas. The betas were pre-adjusted for batch effects using ComBat algorithm. For *cis*-mQTL analysis, all CpG methylation sites up to 100 kb away from EGPA GWAS lead SNP were tested. For *trans*-mQTL analysis, all CpG methylation sites in the seven asthma genes identified by a recent integrative multiomics study^31^ were examined. Briefly, the seven genes (*CHUK, IL1R2, IL1RN, IL5RA, IRF7, LYN,* and *SOCS2*) were found to affect blood cytokine levels, be differentially expressed, and show significant evidence of differential DNA methylation, depending on the asthma status.

### Colocalization analysis

Joint Likelihood Mapping (JLIM version 2.5) was used to test whether the association signals of two traits are driven by a shared genetic effect in a given locus^11^. One of the two compared traits is designated as “primary” and needs to reach the genome-wide significant level of association so that it is safe to assume that there is a true causal variant for the primary trait in the tested locus. The other trait is designated as “secondary” and requires having at least one SNP with the P-value of association < 0.05 with respect to the secondary trait. In the current study, the primary trait GWAS is always the three-way meta-analysis of EGPA combining VCRC, EVGC, and North America II. The secondary traits were either the GWAS of asthma^12^, eosinophil counts^14^, or rheumatoid arthritis^13^, eQTL, or mQTL, depending on the target of colocalization analysis. JLIM was run only on the six genome-wide significant EGPA loci (**Supplementary Table 3**), excluding *HLA-DQ* which is located in the Major Histocompatibility Locus (MHC). The extensive LD in the MHC locus makes a colocalization test challenging due to the reduced genetic resolution. The analysis window of each locus was set to 200 kb centered at the lead SNP for EGPA GWAS peak. In the six loci, JLIM compares the likelihood of observed association signals under three competing hypotheses: 1) there is no causative variant for the secondary trait; 2) the causal variants for primary and secondary traits are identical or in very high LD (r^2^ > 0.8, by default) thus practically indistinguishable; or 3) the causal variants for primary and secondary traits are distinct. The JLIM p-values were calculated by adaptive sampling of null distribution up to 1,000,000 iterations. For GWAS and eQTL data, the association statistics were assumed to follow the correlation structure of a reference panel from the 1000 Genomes Project (Non-Finnish Europeans) since they are all based on populations of European ancestry. For mQTL data, which is based on a Mexican population, we used the in-sample LD of individual-level genetic data. All other parameters for JLIM were set to the default.

#### ACKNOLEDGEMENTS

This research has been conducted using the UK Biobank Resource under Application Number 62142.

## Data Availability

All data produced in the present study are available upon reasonable request to the authors.

---

**Supplementary Figure 1.**
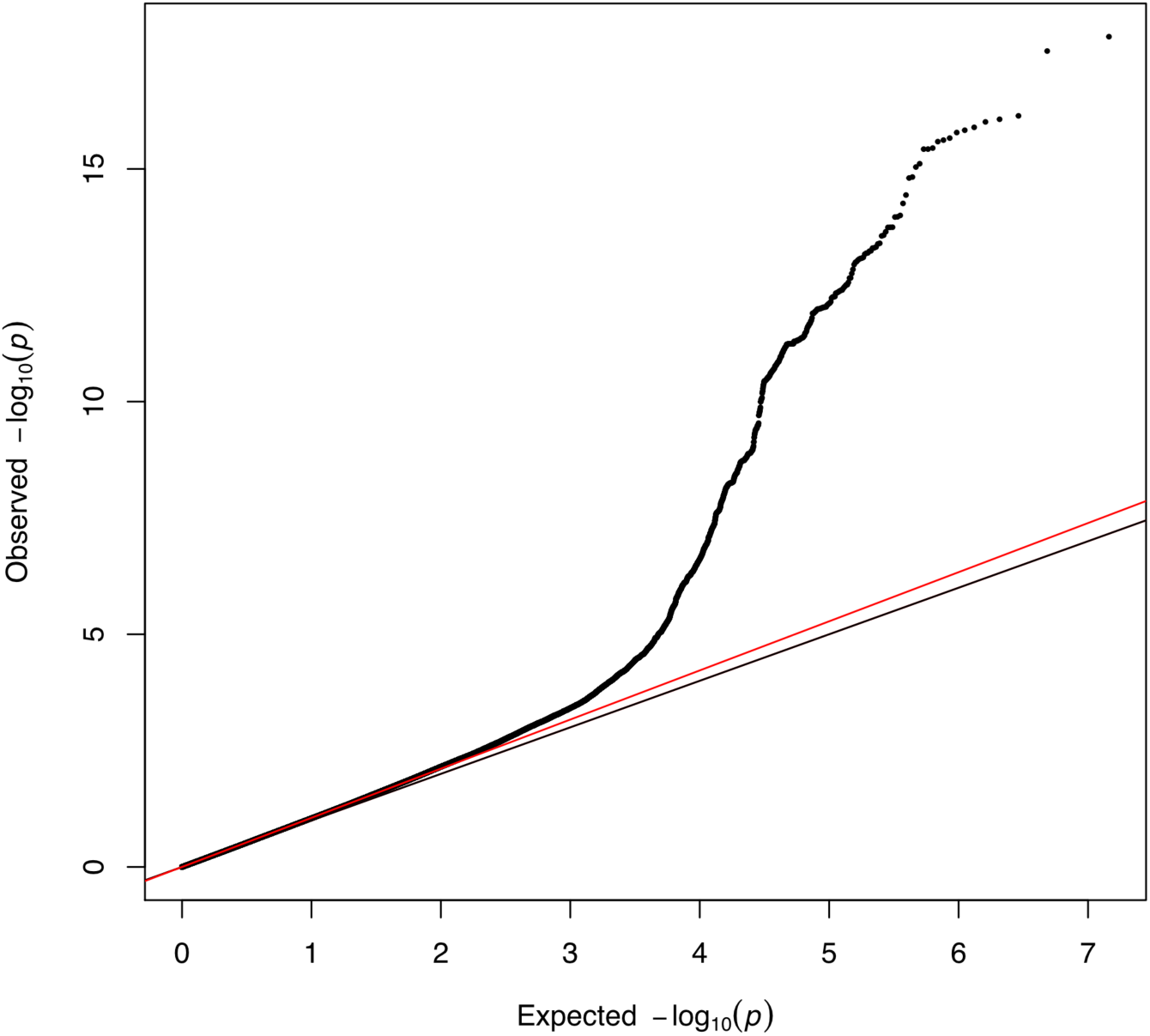
Quantile-Quantile plot of full meta-analysis combining discovery and validation cohorts. Each point is a SNP tested for the association with EGPA. The x axis is the expected p-values under the null. The y axis is the observed p-values. The black line indicates no inflation. The red line indicates the observed level of genomic inflation (α_GC_ = 1.056).

**Supplementary Figure 2.**
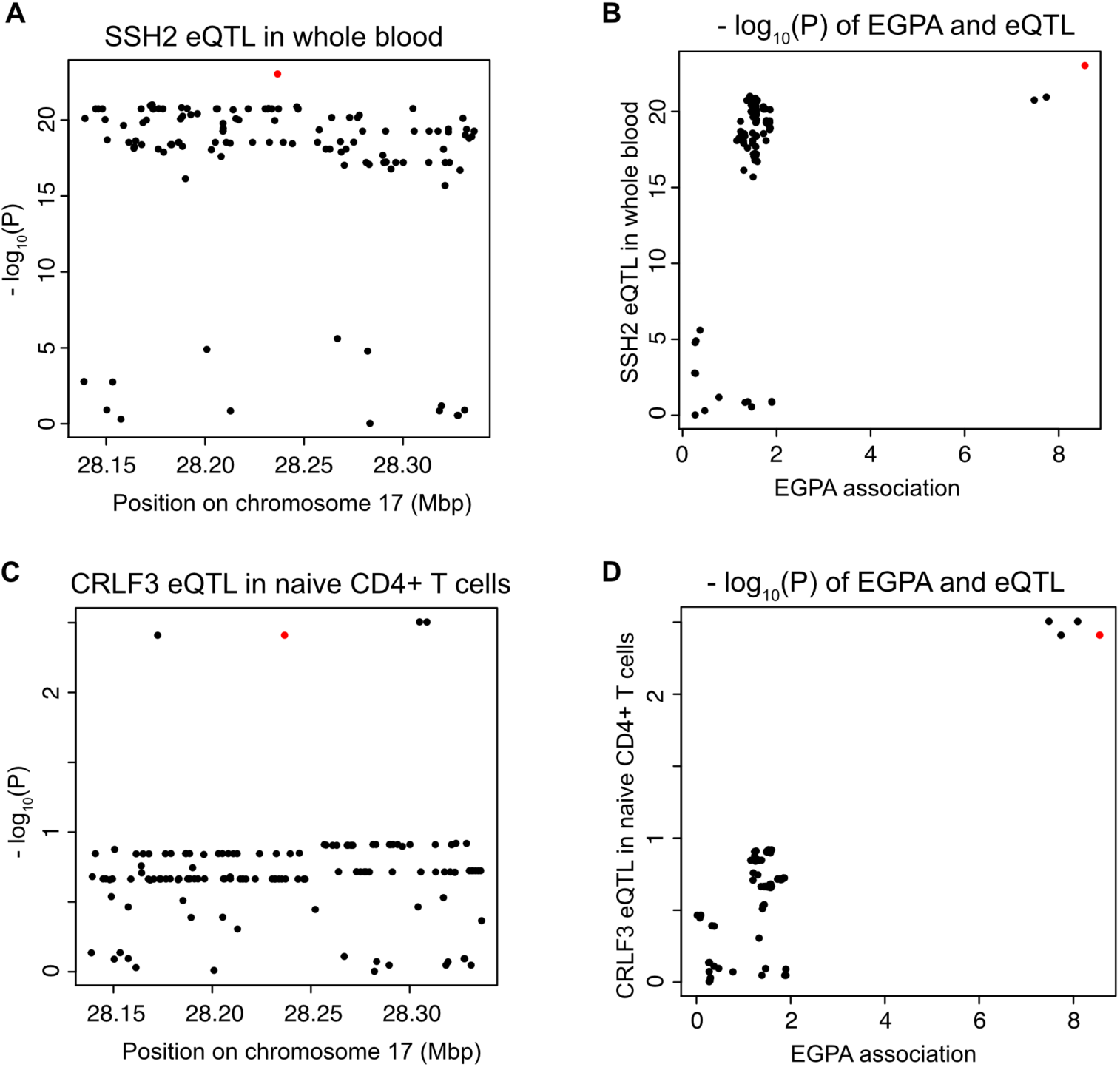
cis-eQTL colocalizing with the EGPA association peak in *SSH2* locus. **(A)** Region plot for *SSH2 cis*-eQTL in whole bloods from Estonian Biobank. **(B)** Scatter plot comparing *SSH2* eQTL association p-values (y axis) with EGPA association p-values (x axis). **(C)** Region plot for *CRLF3 cis*-eQTL in naïve CD4+ T cells from Asthma BRIDGE. **(D)** Scatter plot comparing *CRLF3* eQTL association p-values (y axis) with EGPA association p-values (x axis). Each point represents a SNP. Red point indicates the lead SNP for EGPA association (rs8073292).

**Supplementary Figure 3.**
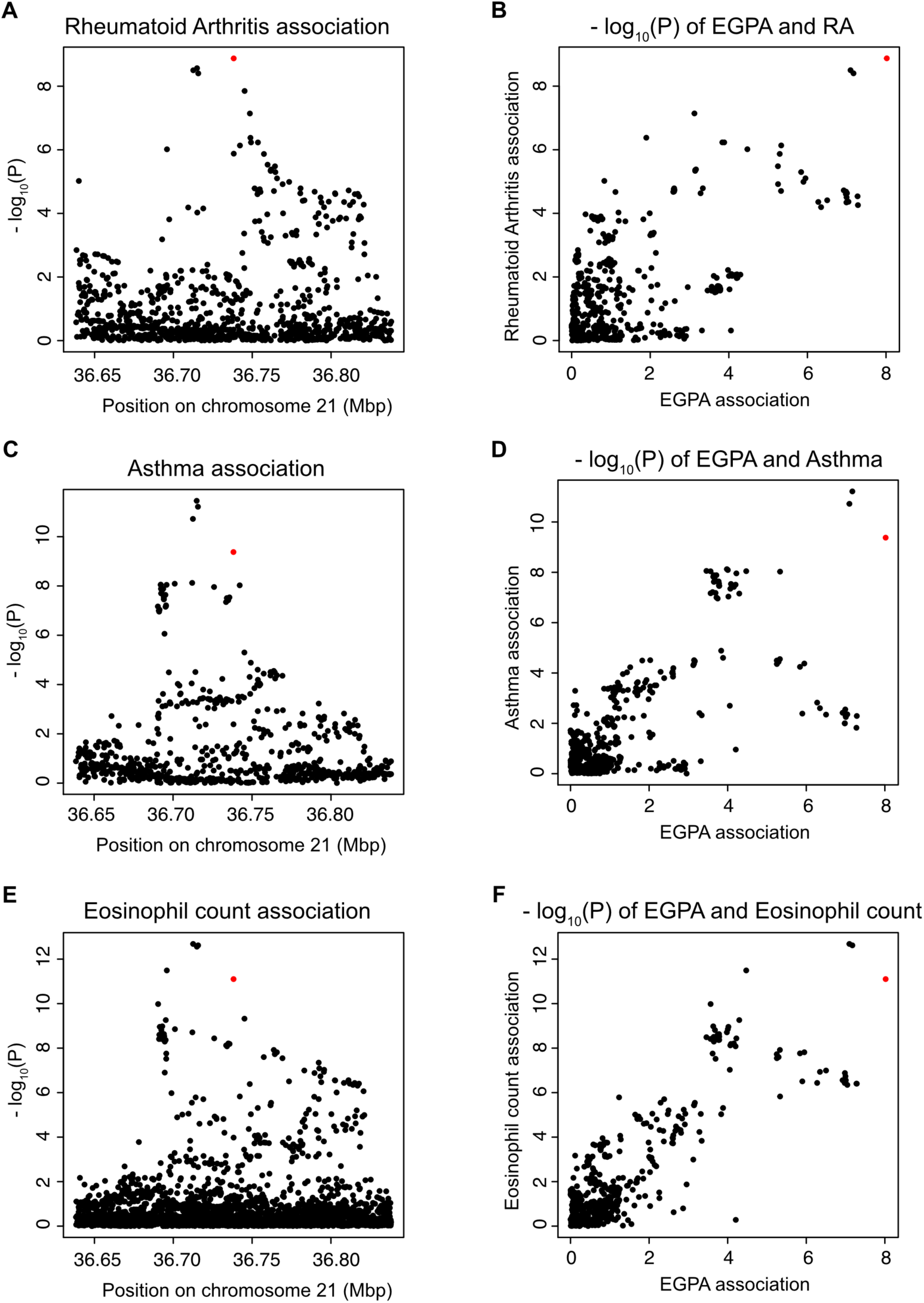
Association peaks of rheumatoid arthritis, asthma, and eosinophil count GWAS colocalizing with the EGPA association peak in *RUNX1* locus. **(A)** Region plot for rheumatoid arthritis (RA) GWAS. **(B)** Scatter plot comparing RA association p-values (y axis) with EGPA association p-values (x axis). **(C)** Region plot for asthma GWAS. **(D)** Scatter plot comparing asthma GWAS (y axis) with EGPA association p-values (x axis). **(E)** Region plot for eosinophil count (EC) GWAS. **(F)** Scatter plot comparing EC GWAS (y axis) with EGPA association p-values (x axis). Each point represents a SNP. Red point indicates the lead SNP for EGPA association (rs8133843).

**Supplementary Figure 4.**
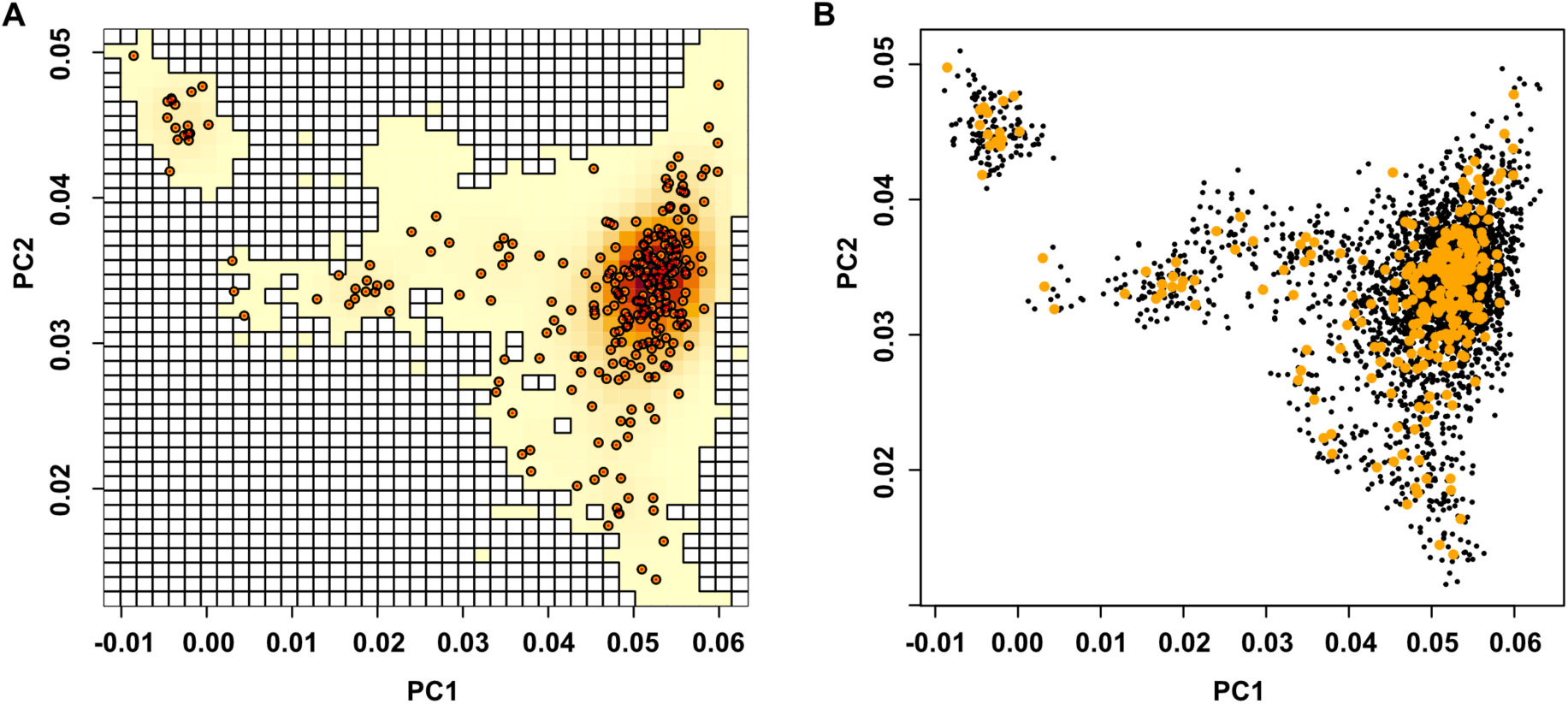
Sampling of control samples from the UK Biobank matching the fine-scale genetic ancestry of VCRC cases. **(A)** The 40 x 40 grid was set up to estimate the kernel density of distribution of VCRC cases in the top two principal component (PC) space. The color of each grid cell represents the density (yellow is the lowest, and red is the highest). White grid cells indicate the masked-out region where UK Biobank samples are too sparse. Orange circles indicate VCRC case subjects. (B) Black dots represent control subjects sampled from the UK Biobank matching the ancestry of VCRC cases (yellow).

**Supplementary Figure 5.**
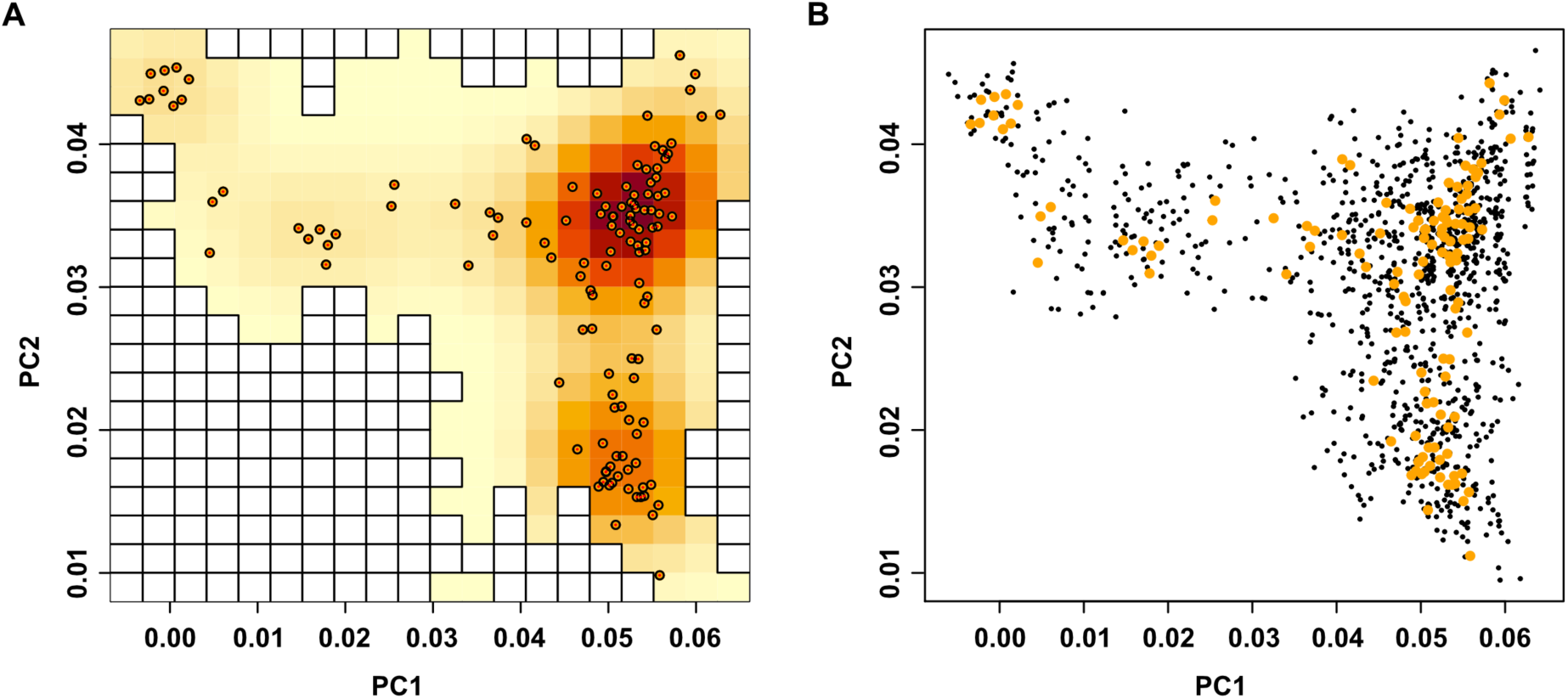
Sampling of control samples from the UK Biobank matching the fine-scale genetic ancestry of North America II cases. **(A)** The 20 x 20 grid was set up to estimate the kernel density of distribution of North America II cases in the top two principal component (PC) space. The color of each grid cell represents the density (yellow is the lowest, and red is the highest). White grid cells indicate the masked-out region where UK Biobank samples are too sparse. Orange circles indicate North America II case subjects. (B) Black dots represent control subjects sampled from the UK Biobank matching the ancestry of North America II cases (yellow).

**Supplementary Table 1.** Replication of previously known EGPA signals in the current study.

| Locus | SNP | Coordinate | EA | EVGC (Lyons et al. 2019) |  |  |  | VCRC (Current Study) |  |  |  |  |  |
| --- | --- | --- | --- | --- | --- | --- | --- | --- | --- | --- | --- | --- | --- |
|  |  |  |  | OR<sub>MPO+</sub> | OR<sub>ACNA-</sub> | OR | P | Proxy (r<sup>2</sup>) | EAF | OR<sub>MPO+</sub> | OR<sub>ACNA-</sub> | OR | P |
| Loci previously identified in all EGPA |  |  |  |  |  |  |  |  |  |  |  |  |  |
| BCL2L11 | rs72946301 | 2:111,861,838 | G | 0.53 | 0.57 | 0.60 | 4.9 x 10<sup>-10</sup> | - | 0.89 | 0.53 | 0.68 | 0.62 | 1.1 x 10<sup>-4</sup> |
| TSLP | rs1837253 | 5:110,401,872 | T | 0.68 | 0.65 | 0.70 | 3.9 x 10<sup>-10</sup> | - | 0.27 | 0.71 | 0.79 | 0.75 | 1.6 x 10<sup>-2</sup> |
| HLA-DQ | rs9274704 | 6:32,637,213 | A | 5.68 | 1.32 | 1.98 | 8.2 x 10<sup>-16</sup> | rs2647025 (0.91) | 0.28 | 2.55 | 1.07 | 1.52 | 2.7 x 10<sup>-5</sup> |
| 10p14 | rs34574566 | 10:9,042,744 | CT | 0.66 | 0.70 | 0.73 | 8.0 x 10<sup>-8</sup> | rs7898135 (0.42) | 0.28 | 0.73 | 0.58 | 0.61 | 7.1 x 10<sup>-6</sup> |
| Loci previously identified in ANCA subtypes |  |  |  |  |  |  |  |  |  |  |  |  |  |
| GPA33 | rs72689399 | 1:167,038,121 | C | 1.12 | 0.19 | 0.37 | 1.1 x 10<sup>-9</sup> (ANCA-) | - | 0.99 | 2.13 | 0.73 | 0.89 | 0.43 (ANCA-) |
| HLA | rs6931740 | 6:32,647,469 | G | 0.55 | 0.61 | 0.62 | 4.2 x 10<sup>-8</sup> (ANCA-) | rs9275109 (0.65) | 0.36 | 0.60 | 0.97 | 0.89 | 0.78 (ANCA-) |
| 12q21 | rs78478398 | 12:80,428,530 | T | 0.17 | 0.81 | 0.59 | 1.7 x 10<sup>-8</sup> (MPO+) | - | 0.97 | 1.08 | 1.35 | 1.18 | 0.87 (MPO+) |
| Loci previously identified by pleiotropy to other traits |  |  |  |  |  |  |  |  |  |  |  |  |  |
| CDK6 | rs42041 | 7:92,246,744 | C | 0.75 | 0.74 | 0.76 | 3.2 x 10<sup>-6</sup> | - | 0.75 | 0.81 | 1.03 | 0.97 | 0.75 |
| IRF1/IL5 | rs11745587 | 5:131,796,922 | G | 0.86 | 0.68 | 0.76 | 4.0 x 10<sup>-7</sup> | - | 0.65 | 0.76 | 0.94 | 0.86 | 0.092 |
| BACH2 | rs6454802 | 6:90,814,199 | C | 1.23 | 1.35 | 1.25 | 3.7 x 10<sup>-6</sup> | - | 0.62 | 1.29 | 1.09 | 1.15 | 0.13 |
| LPP | rs9290877 | 3:188,442,480 | T | 0.68 | 0.81 | 0.79 | 7.7 x 10<sup>-6</sup> | - | 0.68 | 0.73 | 0.97 | 0.89 | 0.22 |

relative to the reference genome GRCh37. Shaded in gray are the ANCA subtype (or the full cohort) where the signal was found significant in the EVGC study.
Significant replication in the current study (Vasculitis Clinical Research Consortium; VCRC) is highlighted in bold. For three loci (*HLA-DQ*, *10p14* and *HLA*), the original
lead SNP is missing in our dataset. Here, we chose closest proxy SNPs in these loci. The proxy SNPs were selected based on the linkage disequilibrium (LD) in Europeans
in the 1000 Genomes Project Phase 3. The sample size for VCRC is n = 295 cases/2898 controls for the full set, n = 81 cases/2898 controls for MPO+ ANCA+ EGPA

**Supplementary Table 2.** Formal replication of eight lead SNPs chosen in discovery meta-analysis.

| Locus | Lead SNP | EA | Discovery |  |  |  |  |  |  |  | Validation |  |  |  |
| --- | --- | --- | --- | --- | --- | --- | --- | --- | --- | --- | --- | --- | --- | --- |
|  |  |  | EVGC |  | VCRC |  |  |  | Discovery meta-analysis<br>(VCRC + EVGC) |  | North America II |  |  |  |
|  |  |  | Beta | P<sup>a</sup> | EAF | INFO | OR | P | Direction<sup>b</sup> | P | EAF | INFO | OR | P |
| *BCL2L11* | 2:111,894,874 | C | -0.04 | $4.9 \times 10^{-10}$ | 0.89 | 0.98 | 0.62 | $1.3 \times 10^{-4}$ | -- | $3.3 \times 10^{-13}$ | 0.87 | 0.96 | 0.68 | $4.3 \times 10^{-2}$ |
| *TSLP* | 5:110,401,872 | T | -0.03 | $3.9 \times 10^{-10}$ | 0.27 | 0.75 | 0.75 | $1.6 \times 10^{-2}$ | -- | $9.5 \times 10^{-11}$ | 0.27 | 0.73 | 0.73 | $1.0 \times 10^{-1}$ |
| *IRF1/IL5* | 5:131,790,616 | G | -0.02 | $2.3 \times 10^{-7}$ | 0.70 | 0.97 | 0.81 | $2.7 \times 10^{-2}$ | -- | $4.2 \times 10^{-8}$ | 0.69 | 1.00 | 0.78 | $8.6 \times 10^{-2}$ |
| *HLA-DQ* | 6:32,635,949 | G | -0.05 | $1.5 \times 10^{-14}$ | 0.72 | 0.91 | 0.66 | $2.9 \times 10^{-5}$ | -- | $4.4 \times 10^{-18}$ | 0.74 | 0.92 | 0.75 | $6.4 \times 10^{-2}$ |
| *OR13F1* | 9:107,266,833 | G | -0.03 | $3.5 \times 10^{-1}$ | 0.98 | 0.92 | 0.16 | $3.3 \times 10^{-15}$ | -- | $5.9 \times 10^{-8}$ | 0.98 | 0.97 | 1.75 | $3.7 \times 10^{-1}$ |
| *10q14* | 10:9,049,253 | C | 0.026 | $5.8 \times 10^{-5}$ | 0.90 | 0.94 | 3.83 | $5.3 \times 10^{-8}$ | ++ | $1.0 \times 10^{-10}$ | 0.91 | 0.88 | 1.92 | $3.1 \times 10^{-2}$ |
| ***SSH2*** | **17:28,105,745** | **G** | **-0.018** | **$6.5 \times 10^{-4}$** | **0.85** | **0.98** | **0.63** | **$3.2 \times 10^{-5}$** | -- | **$1.9 \times 10^{-7}$** | **0.85** | **1** | **0.59** | **$1.9 \times 10^{-3}$** |
| *RUNX1* | 21:36,738,242 | G | -0.021 | $9.6 \times 10^{-7}$ | 0.38 | 0.90 | 0.80 | $2.5 \times 10^{-2}$ | -- | $1.3 \times 10^{-7}$ | 0.40 | 0.95 | 0.71 | $2.2 \times 10^{-2}$ |

Coordinates are relative to the reference genome GRCh37. The lead SNPs were chosen by discovery meta-analysis p-values $< 5 \times 10^{-7}$ . Discovery meta-analysis combines the association statistics of EVGC and VCRC in a summary statistics level. Significant formal replication in North America II are marked in bold ( $P < 6.25 \times 10^{-3}$ after the multiple testing correction of 8 SNPs). **a.** The EVGC p-values were adjusted by $\lambda_{GC}$ to account for $\lambda_{GC} = 1.047$ in EVGC GWAS data. **b.** Direction of effect in EVGC and VCRC. EA = Effect Allele. EAF = Effect Allele Frequency. OR = Odds Ratio.

**Supplementary Table 3.** Genome-wide significant loci in three-way meta-analysis across EVGC, VCRC and North America II.

|  |  |  | Discovery |  |  |  |  |  |  | Validation |  |  |  | Full three-way meta-analysis (D+V) |
| --- | --- | --- | --- | --- | --- | --- | --- | --- | --- | --- | --- | --- | --- | --- |
|  |  |  | EVGC |  | VCRC |  |  |  | North America II |  |  |  |  |  |
| Locus | Lead SNP | EA | Beta | P<sup>a</sup> | EAF | INFO | OR | P | EAF | INFO | OR | P | Direction<sup>b</sup> | P |
| *BCL2L11* | 2:111,900,560 | T | -0.04 | 6.1 x 10<sup>-10</sup> | 0.89 | 0.98 | 0.62 | 1.1 x 10<sup>-4</sup> | 0.88 | 0.97 | 0.67 | 3.6 x 10<sup>-2</sup> | --- | 4.1 x 10<sup>-14</sup> |
| *TSLP* | 5:110,401,872 | T | -0.03 | 3.9 x 10<sup>-10</sup> | 0.27 | 0.75 | 0.75 | 1.6 x 10<sup>-2</sup> | 0.27 | 0.73 | 0.73 | 1.0 x 10<sup>-1</sup> | --- | 2.8 x 10<sup>-11</sup> |
| *IRF1/IL5* | 5:131,797,547 | A | -0.02 | 5.0 x 10<sup>-7</sup> | 0.61 | 0.96 | 0.83 | 3.4 x 10<sup>-2</sup> | 0.60 | 0.98 | 0.67 | 3.9 x 10<sup>-3</sup> | --- | 2.5 x 10<sup>-9</sup> |
| *HLA-DQ* | 6:32,635,949 | G | -0.05 | 1.5 x 10<sup>-14</sup> | 0.72 | 0.91 | 0.66 | 2.9 x 10<sup>-5</sup> | 0.74 | 0.92 | 0.75 | 6.4 x 10<sup>-2</sup> | --- | 1.4 x 10<sup>-18</sup> |
| *10q14* | 10:9,049,253 | C | 0.026 | 5.8 x 10<sup>-5</sup> | 0.90 | 0.94 | 3.83 | 5.3 x 10<sup>-8</sup> | 0.91 | 0.88 | 1.92 | 3.1 x 10<sup>-2</sup> | +++ | 9.6 x 10<sup>-12</sup> |
| *SSH2* | 17:28,236,645 | A | 0.018 | 6.7 x 10<sup>-4</sup> | 0.17 | 0.86 | 1.58 | 5.3 x 10<sup>-5</sup> | 0.16 | 0.93 | 1.76 | 9.0 x 10<sup>-4</sup> | +++ | 2.8 x 10<sup>-9</sup> |
| *RUNX1* | 21:36,738,242 | G | -0.021 | 9.6 x 10<sup>-7</sup> | 0.38 | 0.90 | 0.80 | 2.5 x 10<sup>-2</sup> | 0.40 | 0.95 | 0.71 | 2.2 x 10<sup>-2</sup> | --- | 9.5 x 10<sup>-9</sup> |

Coordinates are relative to the reference genome GRCh37. Genome-wide significant loci are based on three-way meta-analysis p-value $< 5 \times 10^{-8}$ .
Three-way meta-analysis combines the association statistics across EVGC, VCRC and North America II in a summary statistics level. **a.** EVGC p-values were adjusted by $\lambda_{GC}$ to account for $\lambda_{GC} = 1.047$ in EVGC GWAS data. **b.** Direction of effect in EVGC, VCRC and North America II. EA = Effect Allele. EAF = Effect Allele Frequency. OR = Odds Ratio.

**Supplementary Table 4.**
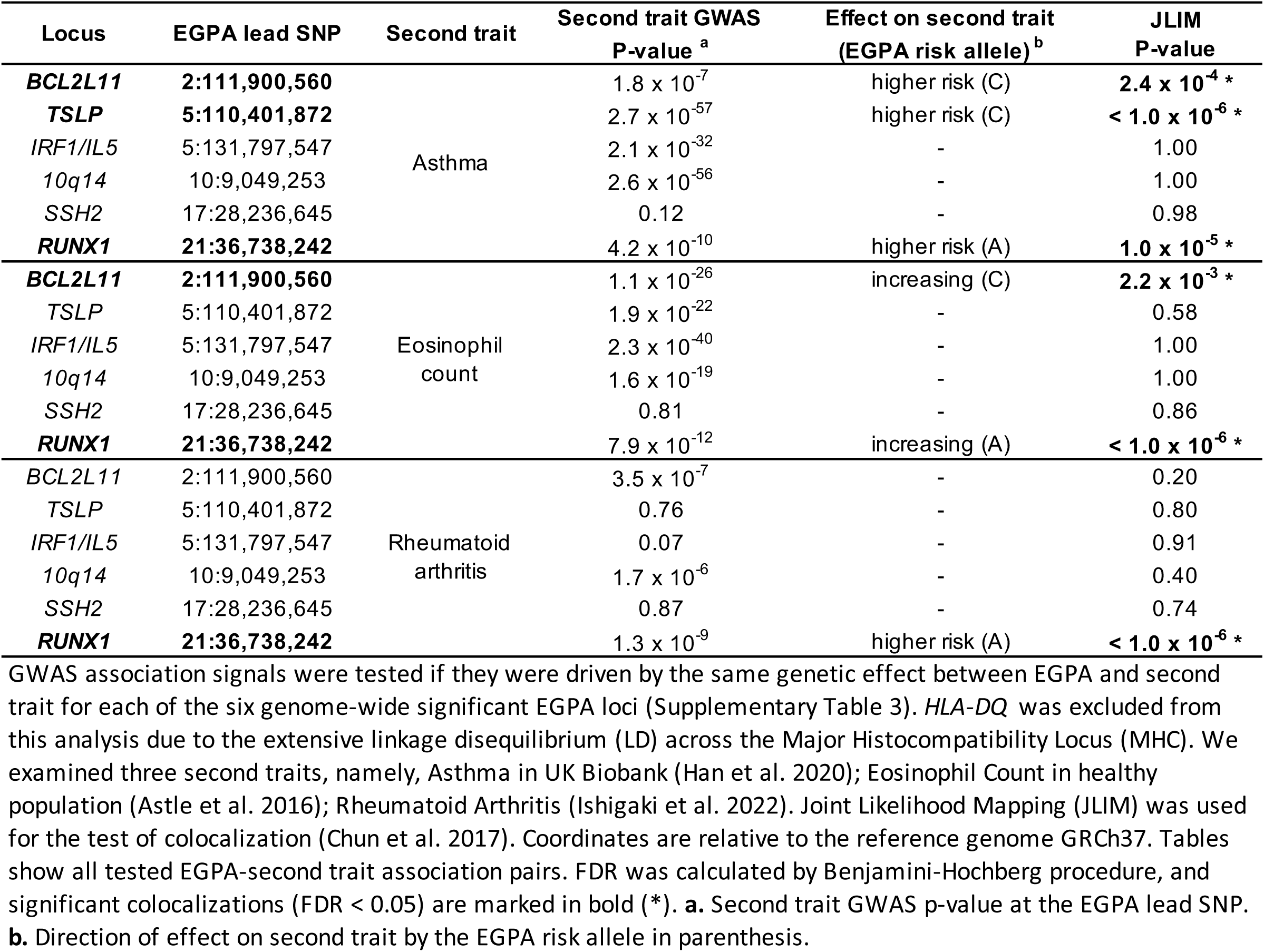
Pleiotropy between EGPA and other traits in EGPA-associated loci (FDR < 0.05).

**Supplementary Table 5.**
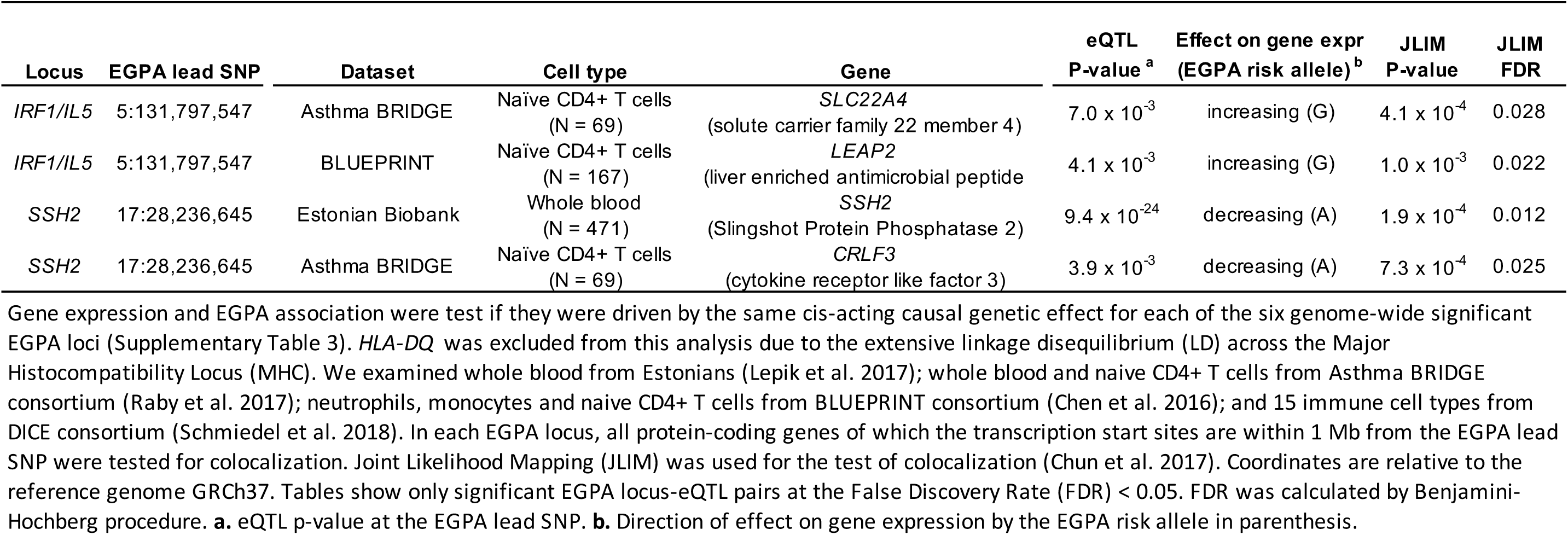
cis-eQTL colocalizing with EGPA association peaks (FDR < 0.05).

| Locus | EGPA lead SNP | Dataset | Cell type | Gene | eQTL<br>P-value<sup>a</sup> | Effect on gene expr<br>(EGPA risk allele)<sup>b</sup> | JLIM<br>P-value | JLIM<br>FDR |
| --- | --- | --- | --- | --- | --- | --- | --- | --- |
| *IRF1/IL5* | 5:131,797,547 | Asthma BRIDGE | Naïve CD4+ T cells<br>(N = 69) | *SLC22A4*<br>(solute carrier family 22 member 4) | $7.0 \times 10^{-3}$ | increasing (G) | $4.1 \times 10^{-4}$ | 0.028 |
| *IRF1/IL5* | 5:131,797,547 | BLUEPRINT | Naïve CD4+ T cells<br>(N = 167) | *LEAP2*<br>(liver enriched antimicrobial peptide) | $4.1 \times 10^{-3}$ | increasing (G) | $1.0 \times 10^{-3}$ | 0.022 |
| *SSH2* | 17:28,236,645 | Estonian Biobank | Whole blood<br>(N = 471) | *SSH2*<br>(Slingshot Protein Phosphatase 2) | $9.4 \times 10^{-24}$ | decreasing (A) | $1.9 \times 10^{-4}$ | 0.012 |
| *SSH2* | 17:28,236,645 | Asthma BRIDGE | Naïve CD4+ T cells<br>(N = 69) | *CRLF3*<br>(cytokine receptor like factor 3) | $3.9 \times 10^{-3}$ | decreasing (A) | $7.3 \times 10^{-4}$ | 0.025 |

**Supplementary Table 6.**
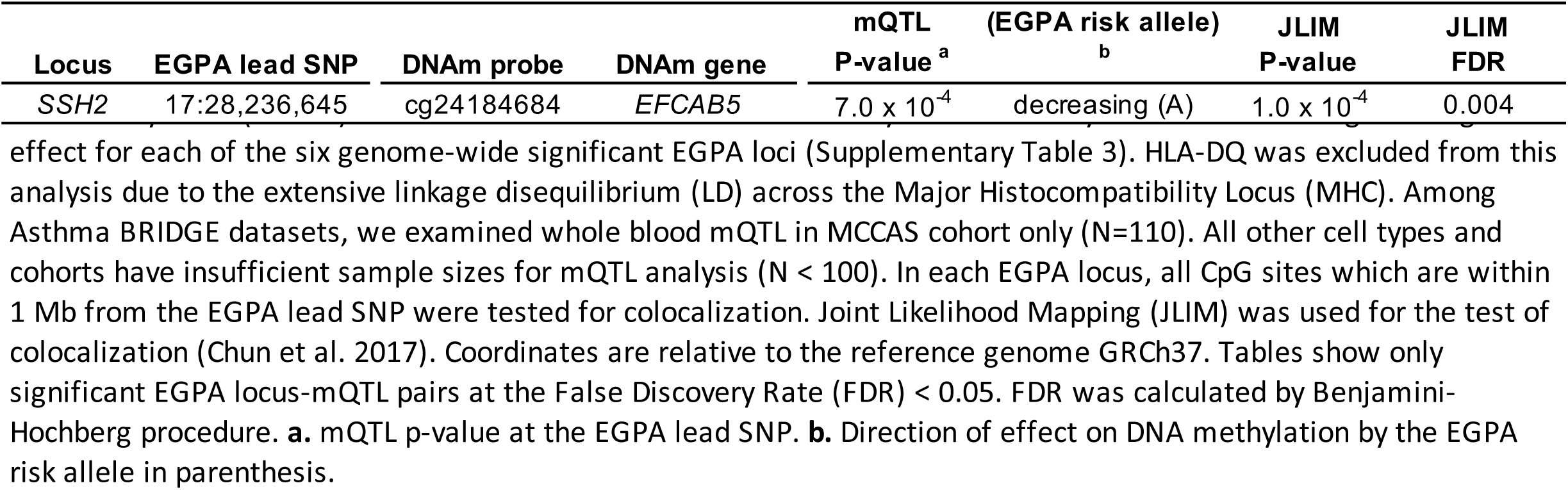
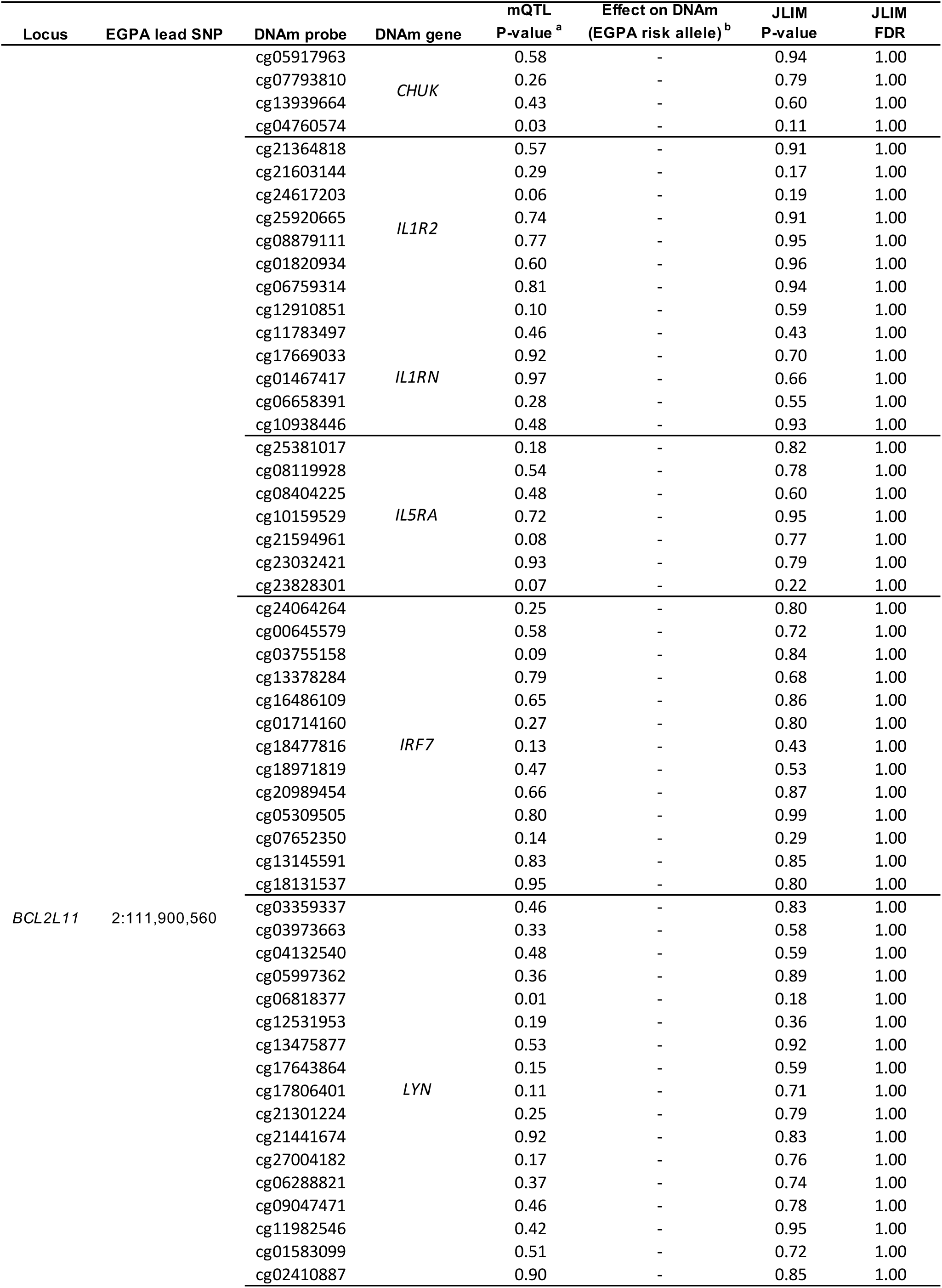
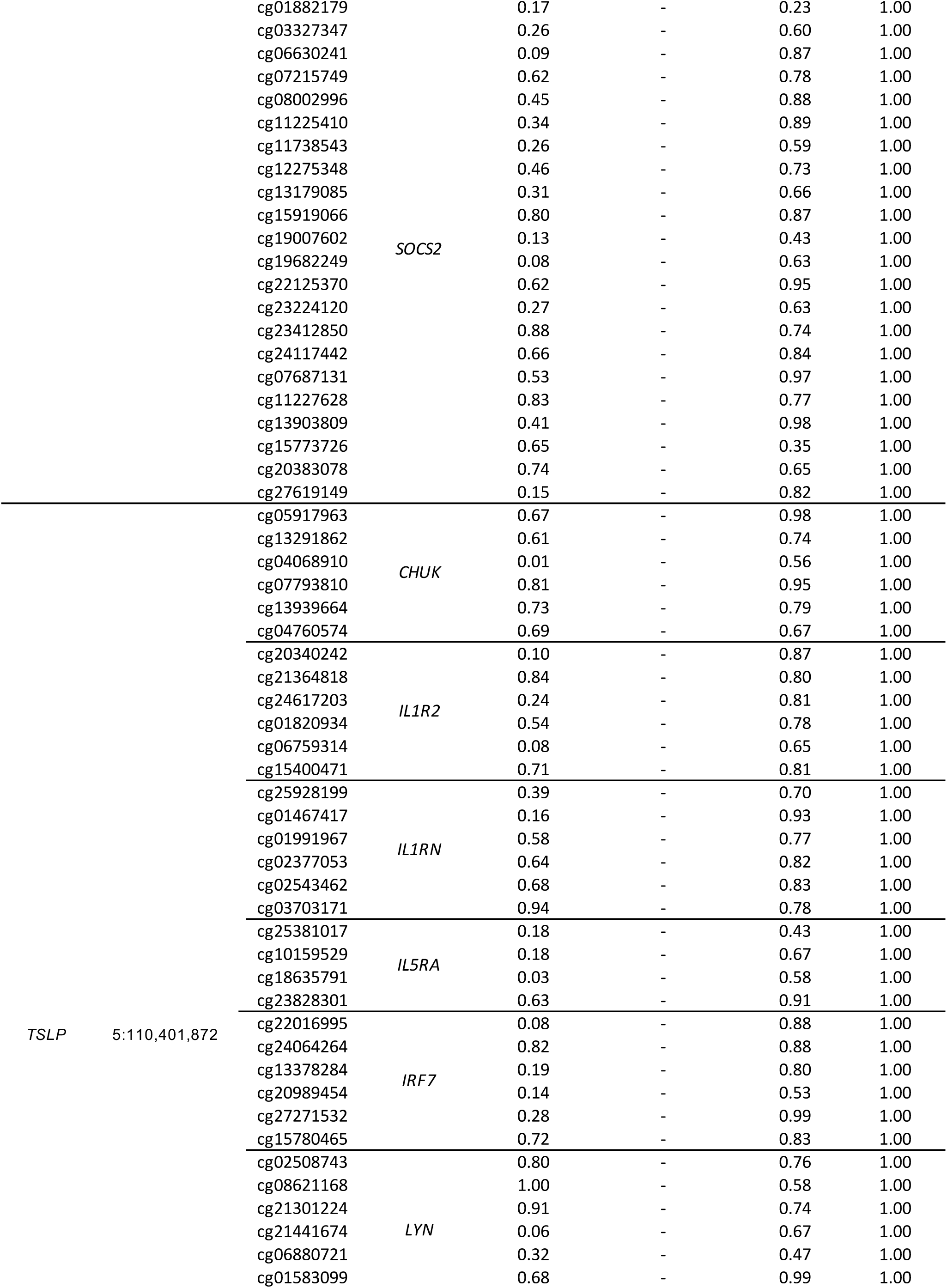
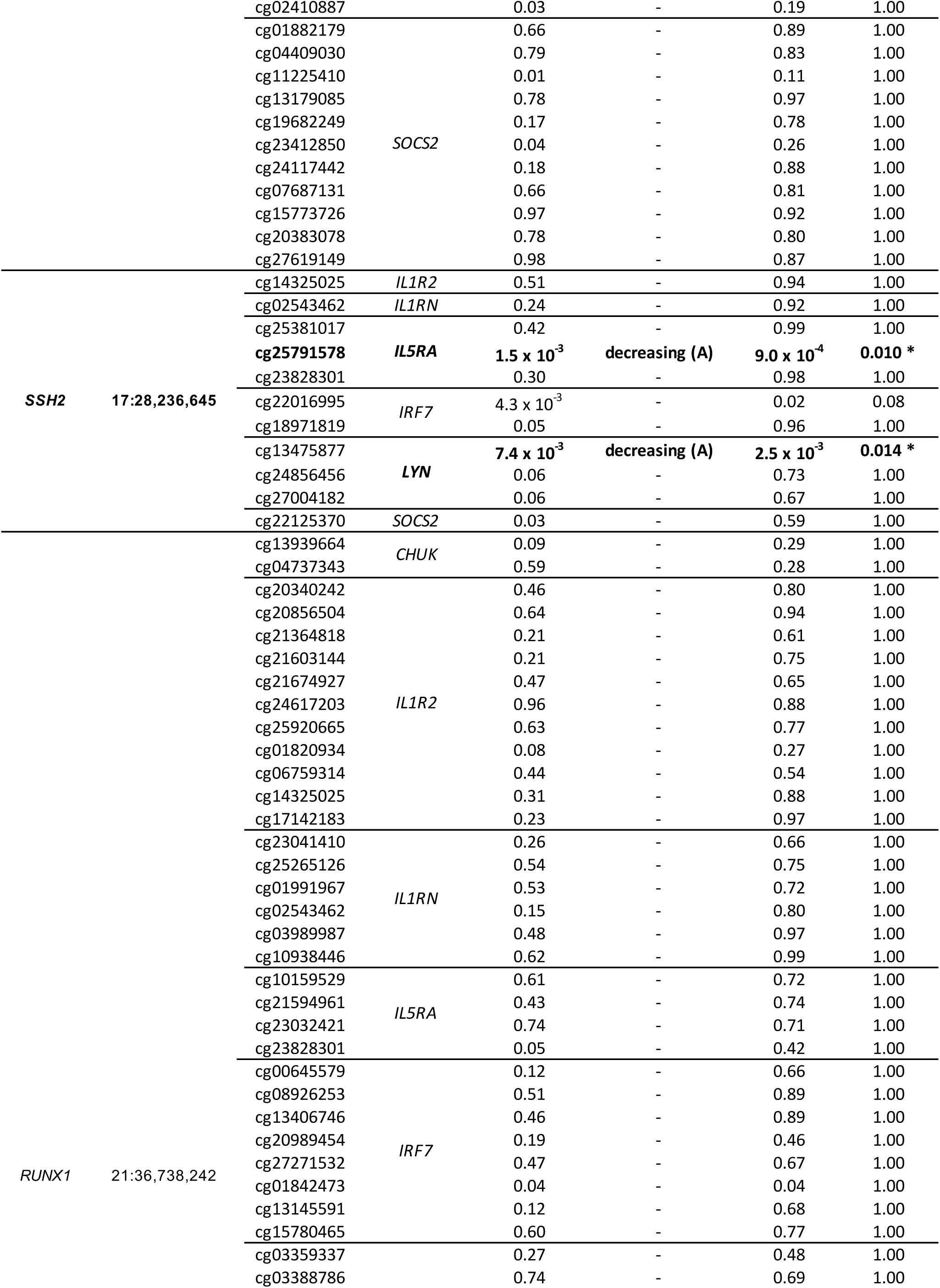
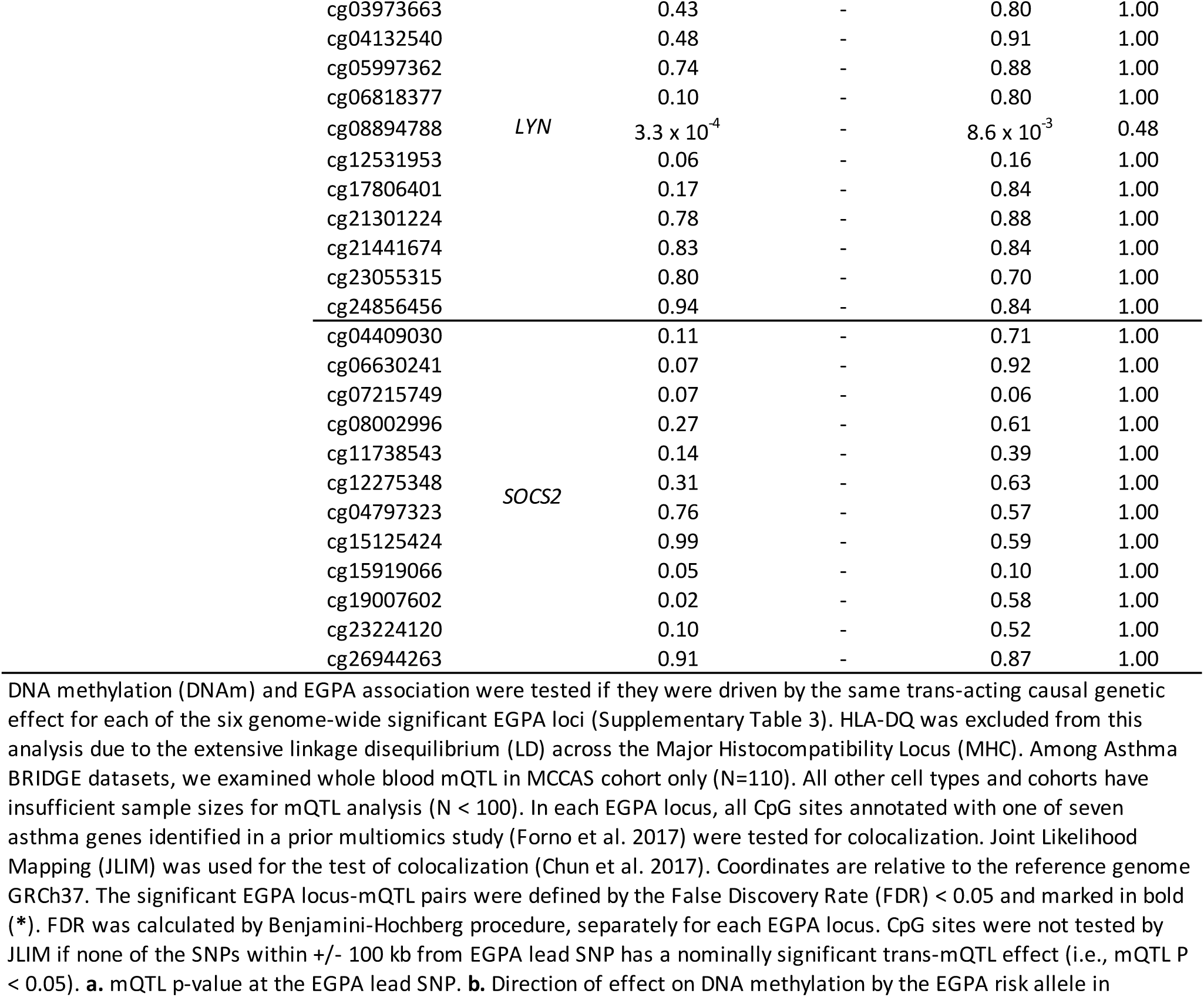
cis-mQTL colocalizing with EGPA association peaks in Asthmatics (FDR < 0.05).

**Supplementary Table 8.**
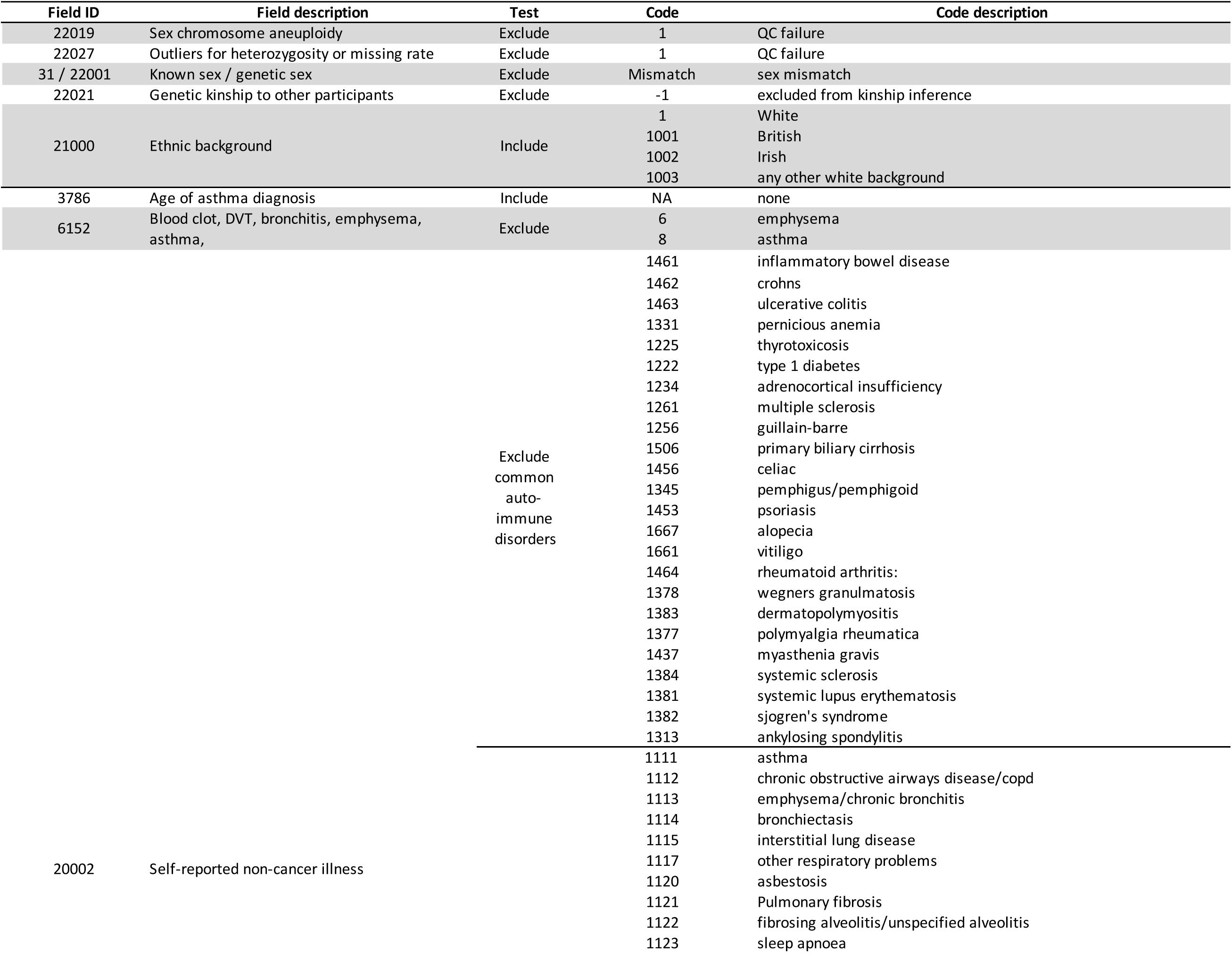
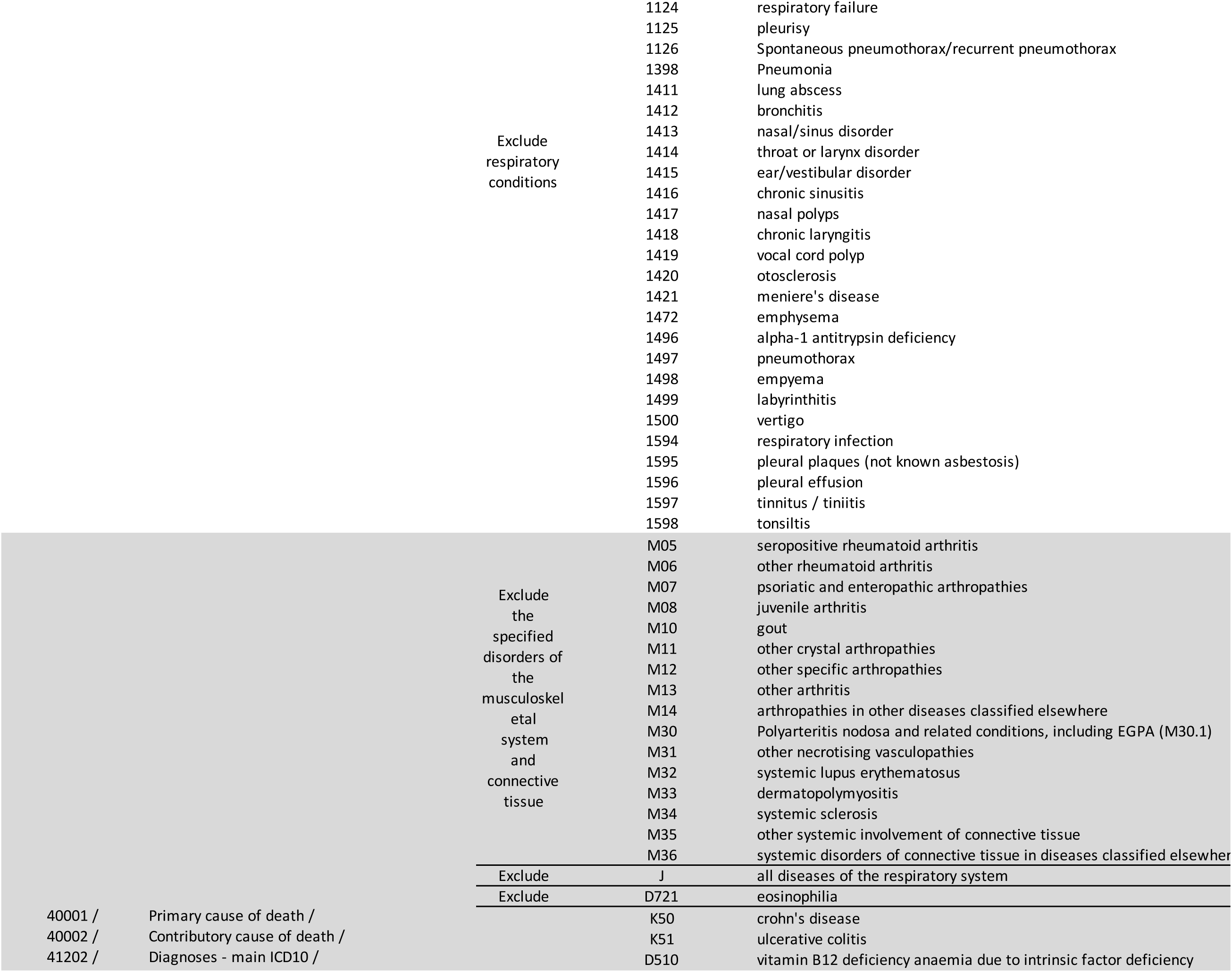
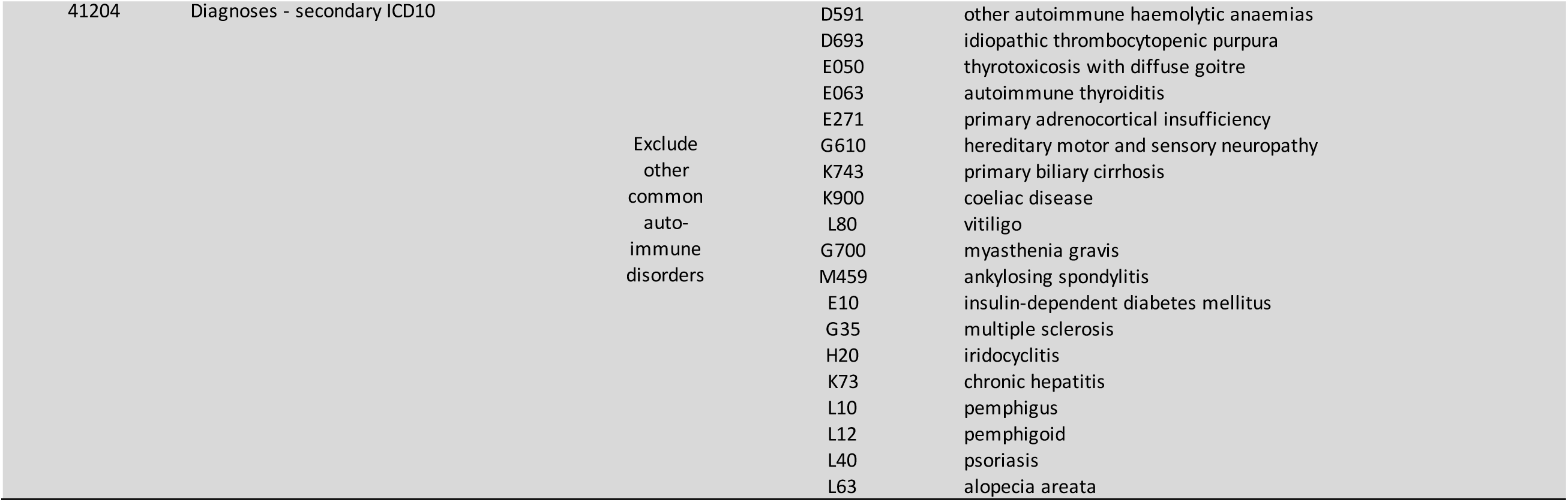
Exclusion criteria for control subjects.

